# Immunomics analysis of rheumatoid arthritis identified precursor dendritic cells as a key cell subset of treatment resistance

**DOI:** 10.1101/2022.12.21.22283652

**Authors:** Saeko Yamada, Yasuo Nagafuchi, Min Wang, Mineto Ota, Hiroaki Hatano, Yusuke Takeshima, Mai Okubo, Satomi Kobayashi, Yusuke Sugimori, Masahiro Nakano, Ryochi Yoshida, Norio Hanata, Yuichi Suwa, Yumi Tsuchida, Yukiko Iwasaki, Shuji Sumitomo, Kanae Kubo, Kenichi Shimane, Keigo Setoguchi, Takanori Azuma, Hiroko Kanda, Hirofumi Shoda, Xuan Zhang, Kazuhiko Yamamoto, Kazuyoshi Ishigaki, Tomohisa Okamura, Keishi Fujio

**Affiliations:** Department of Allergy and Rheumatology, Graduate School of Medicine, the University of Tokyo, Tokyo, Japan; Department of Functional Genomics and Immunological Diseases, Graduate School of Medicine, the University of Tokyo, Tokyo, Japan; Department of Rheumatology, Beijing Hospital, National Center of Gerontology, Institute of Geriatric Medicine, Clinical Immunology Center, Graduate School of Peking Union Medical College, Chinese Academy of Medical Sciences and Peking Union Medical College, Beijing, China; Laboratory for Human Immunogenetics, Center for Integrative Medical Sciences, RIKEN, Kanagawa, Japan; Department of Medicine and Rheumatology, Tokyo Metropolitan Geriatric Hospital, Tokyo, Japan; Department of Rheumatology, Tokyo Metropolitan Bokutoh Hospital, Tokyo, Japan; Laboratory for Autoimmune Diseases, Center for Integrative Medical Sciences, RIKEN, Kanagawa, Japan; Allergy and Immunological Diseases, Tokyo Metropolitan Cancer and Infectious Diseases Center Komagome Hospital, Tokyo, Japan; Azuma Rheumatology Clinic, Saitama, Japan; Immune-Mediated Diseases Therapy Center, Graduate School of Medicine, The University of Tokyo, Tokyo, Japan

**Author notes:** **Correspondence** Dr. Yasuo Nagafuchi MD. PhD., Department of Allergy and Rheumatology and Department of Functional Genomics and Immunological, Diseases, Graduate School of Medicine, The University of Tokyo, 7-3-1 Hongo, Bunkyo-ku, Tokyo, 113-8655, Japan, E-mail address, Dr. Keishi Fujio MD. PhD., Department of Allergy and Rheumatology, Graduate School of Medicine, The University of Tokyo, 7-3-1 Hongo, Bunkyo-ku, Tokyo, 113-8655, Japan. SY and YN contributed equally.

**Keywords:** rheumatoid arthritis, treatment resistance, predictors, immunomics, dendritic cells precursors

## Abstract

**Objectives:** Little is known about the immunology underlying variable treatment response in rheumatoid arthritis (RA). We performed large scale transcriptome analyses of peripheral blood immune cell subsets to identify immune cells that predict treatment resistance.

**Methods:** We isolated 18 peripheral blood immune cell subsets of 55 pre-treatment RA patients and 39 healthy controls, and performed RNA sequencing. Transcriptome changes in RA and treatment effects were systematically characterized. Association between immune cell gene modules and treatment resistance was evaluated. We validated predictive value of identified parameters for treatment resistance using quantitative polymerase chain reaction (qPCR) and mass cytometric analysis cohorts. We also characterized the identified population by synovial single cell RNA-seq analysis.

**Results:** Immune cells of RA patients were characterized by enhanced interferon and IL6-JAK-STAT3 signaling that demonstrate partial normalization after treatment. A gene expression module of plasmacytoid dendritic cells (pDC) reflecting the expansion of pre-dendritic cells (pre-DC) exhibited strongest association with treatment resistance. Type I interferon signaling was negatively correlated to pre-DC gene expression. qPCR and mass cytometric analysis in independent cohorts validated that the pre-DC associated gene expression and the proportion of pre-DC were significantly higher before treatment in treatment-resistant patients. A cluster of synovial DCs showed both features of pre-DC and proinflammatory conventional DC2s.

**Conclusions:** An increase in pre-DC in peripheral blood predicted RA treatment resistance. Pre-DC could have pathophysiological relevance to RA treatment response.

**Key messages:** *What is already known about this subject?:*

- Limited information is available about the immune cells that are associated with RA treatment resistance.

*What does this study add?:*

- RA treatment resistance can be predicted by an increase in pre-DC in peripheral blood prior to treatment.
- The expression of genes reflecting an increase in pre-DC is negatively correlated to the type I interferon signature, which is associated with good therapeutic response.
- Synovial pre-DC-like cells are proinflammatory cDC2s.

*How might this impact on clinical practice or future developments?:*

- Stratified treatment of RA is possible using pre-DC as a biomarker, and it might be possible to develop new therapies for treatment-resistant RA by targeting pre-DC.

## Introduction

Rheumatoid arthritis (RA) is a common chronic autoimmune inflammatory disease characterized by persistent synovitis and/or joint destruction. Difficult-to-treat (D2T) RA is recognized as an unsolved problem in the clinical setting (1-4). In the treatment of RA, both patients and healthcare providers need precision medicine that stratifies treatment based on the predictions of treatment response.

To elucidate the causes of RA and predict therapeutic efficacy, transcriptome analyses have been performed on peripheral blood mononuclear cells (PBMC) (5, 6) and synovia (7, 8). To date, type I interferon (IFN) gene signature or activity (5, 9, 10) has been proposed as predictive factor of treatment response. As for myeloid cells, the evidence suggests a relationship between treatment response and myeloid cells including dendritic cells (DC). An increase in conventional (c) DC in peripheral blood is correlated with treatment response to a tumor necrosis factor (TNF) inhibitor, infliximab (IFX) (11). For adaptive immune cells, T cells (12-14) and B cells (15-18) also exhibited association with therapeutic response. In RA synovium, treatment response showed association with expression of DC adhesion molecules (19). Synovial macrophage and myeloid DC gene signatures were associated with higher response rates to interleukin (IL)-6 receptor inhibitor, tocilizumab (TCZ) (8).

Dendritic cell precursors (pre-DC) are recently identified subpopulation of DCs. In 2017, See P *et al*. demonstrated that there are cells that are included in the subset of cells that have conventionally been considered plasmacytoid (p) DC, and that while pre-DC shares many of the same markers as pDC, they differentiate into cDC1 and cDC2 (20). They propose that IL-12 production and naive CD4^+^ T cell stimulation, which have traditionally been considered functions of pDC, are in fact functions of pre-DC, not pDC. Compared to pDC, pre-DC characteristically express *CD33 (SIGLEC3), CX3CR1, SIGLEC6 (CD327), CD2*, and *CD5*. In 2017, Villani AC *et al*. also identified DC subfractions DC1 through DC6 by means of single cell (sc) RNA-seq of human peripheral blood, and identified AS DC as a DC subfraction that was characterized by the expression of *AXL, SIGLEC1*, and *SIGLEC6* and that powerfully activated T cells (21). It is believed that pre-DC and AS DC are largely overlapping populations (22).

To date, there have been no comprehensive studies of the immune cells that play a pivotal role in treatment-resistant RA. We recently constructed an atlas for the detailed gene expression profiles of peripheral blood immune cells in patients with immune diseases (the Immune Cell Gene Expression Atlas from the University of Tokyo, or ImmuNexUT) (23). In this study, we performed a comprehensive assessment of the transcriptome profiles of immune cells in peripheral blood prior to treatment in a total of 55 patients with RA, 24 of whom had been reported in ImmuNexUT and 31 of whom were newly added, and assessed the gene expressions and subsets that predict treatment resistance. Then, we evaluated the association of pre-DC with treatment-resistance of RA in two independent cohorts.

## Materials and Methods

**See online supplementary materials and methods**.

## Results

### RA and HC clinical findings

To identify parameters related to the response to molecular targeted therapy, we recruited 55 active RA patients who required addition or switching of disease modifying anti rheumatic drugs (DMARDs) (online supplementary method). We also included 39 healthy control (HC) volunteers in the analysis (figure 1A). No significant differences in the age or sex were found between the RA and HC populations (online supplementary table 1).

**Figure 1.**
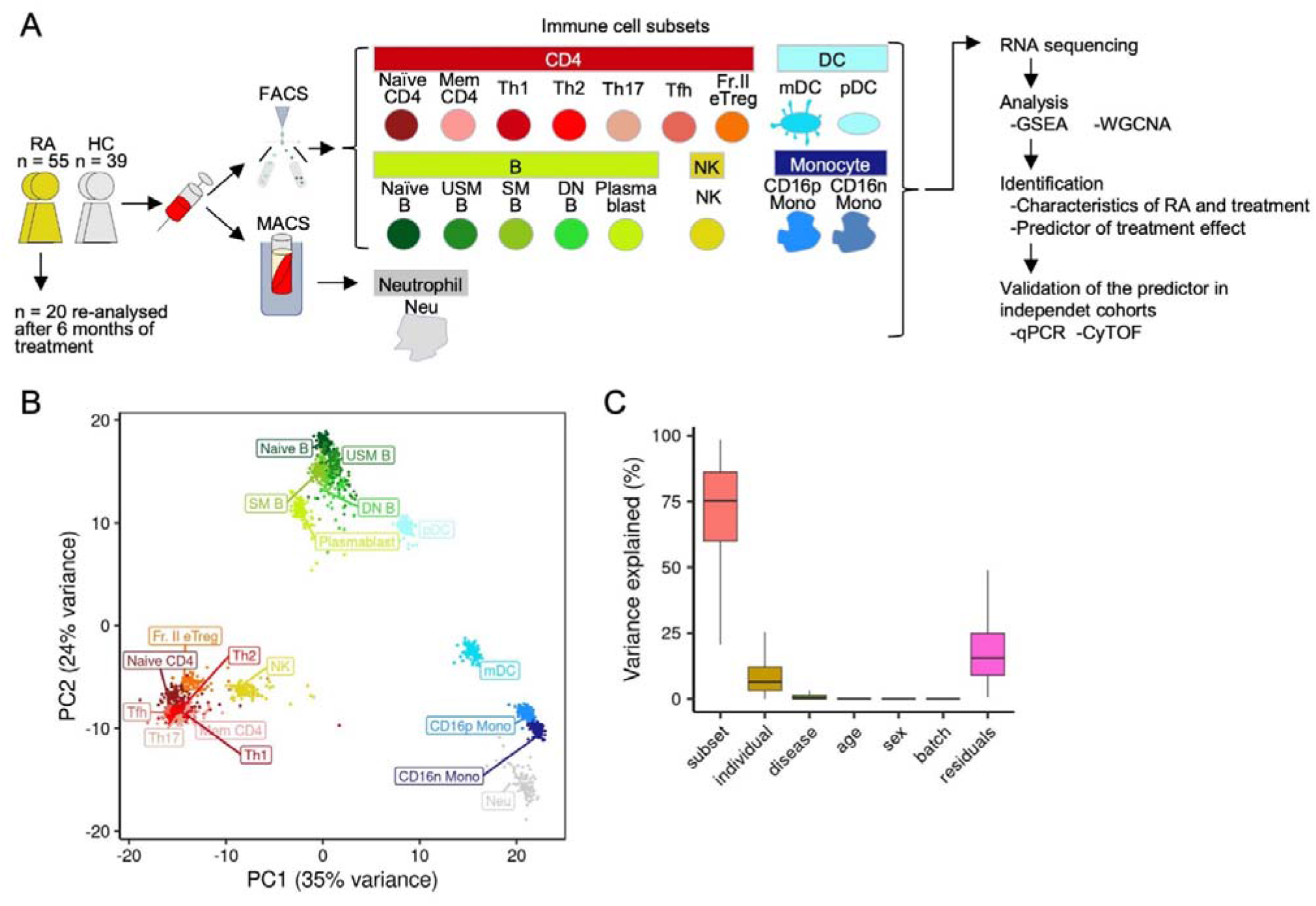
Overview of the study. (A) Study concept. (B) Using all RNA-seq samples from the RA (n = 55) (before treatment/after treatment) and HC (n = 39) populations, PCA was performed using the 500 most highly variable genes. (C) A linear mixed model was used to perform gene expression variance decomposition on the RNA-seq samples. The fixed effect or age on gene expression, and the random effects of the immune cell subset, individual, difference between RA and HC (disease), sex, and each of the 4 dataset batches was calculated. RA: rheumatoid arthritis, HC: healthy control, FACS: fluorescence-activated cell sorting, MACS: magnetic-activated cell sorting, GSEA: gene set enrichment analysis, WGCNA: weighted gene co-expression network analysis, qPCR: quantitative polymerase chain reaction, CyTOF: cytometry by time of flight, PCA: principal component analysis. Definitions of the subsets are presented in online supplementary table 2.

### Overall picture of RNA-seq analysis by immune cell subset

We performed RNA-seq for 55 RA patients and 39 HC volunteers for each of the 18 peripheral blood immune cell subsets, sorted or isolated based on cell surface antigens (online supplementary table 2). We performed a second RNA-seq analysis for 20 (36.4%) of the 55 patients (of these 20, 15 received abatacept (ABT) and 5 TCZ) at 6 months after treatment initiation to assess effects of treatment on immune system using gene expression. After stringent quality control, 1,701 samples with consistent gene expression pattern were included in the analysis (online supplementary method, online supplementary figure 1A). In a principal component analysis (PCA), gene expression profiles of the different samples in each subset were similar, with only minor differences between the RA and HC populations (figure 1B, online supplementary figure 1B). Most of the variance in gene expression was explained by the subset (median 74%) and the individual (median 6.1%), though some (median 0.45%) depended on the differences between the RA and HC populations (figure 1C). The explained variance associated with the batches between the datasets was only 0.00055% (median), demonstrating the appropriateness of combining the datasets (online supplementary method).

### Genes with varying expression in RA immune cells and therapeutic medication efficacy

For investigating the differences in each type of immune cell between the RA and HC populations, we performed a Gene Set Enrichment Analysis (GSEA) for each subset of genes with varying expression in the RA population prior to treatment relative to the HC population (figure 2A). Increased expression of IFN response genes and IL6-Janus kinase (JAK)-signal transducer and activator of transcription (STAT) response genes was found in various subsets with RA population.

**Figure 2.**
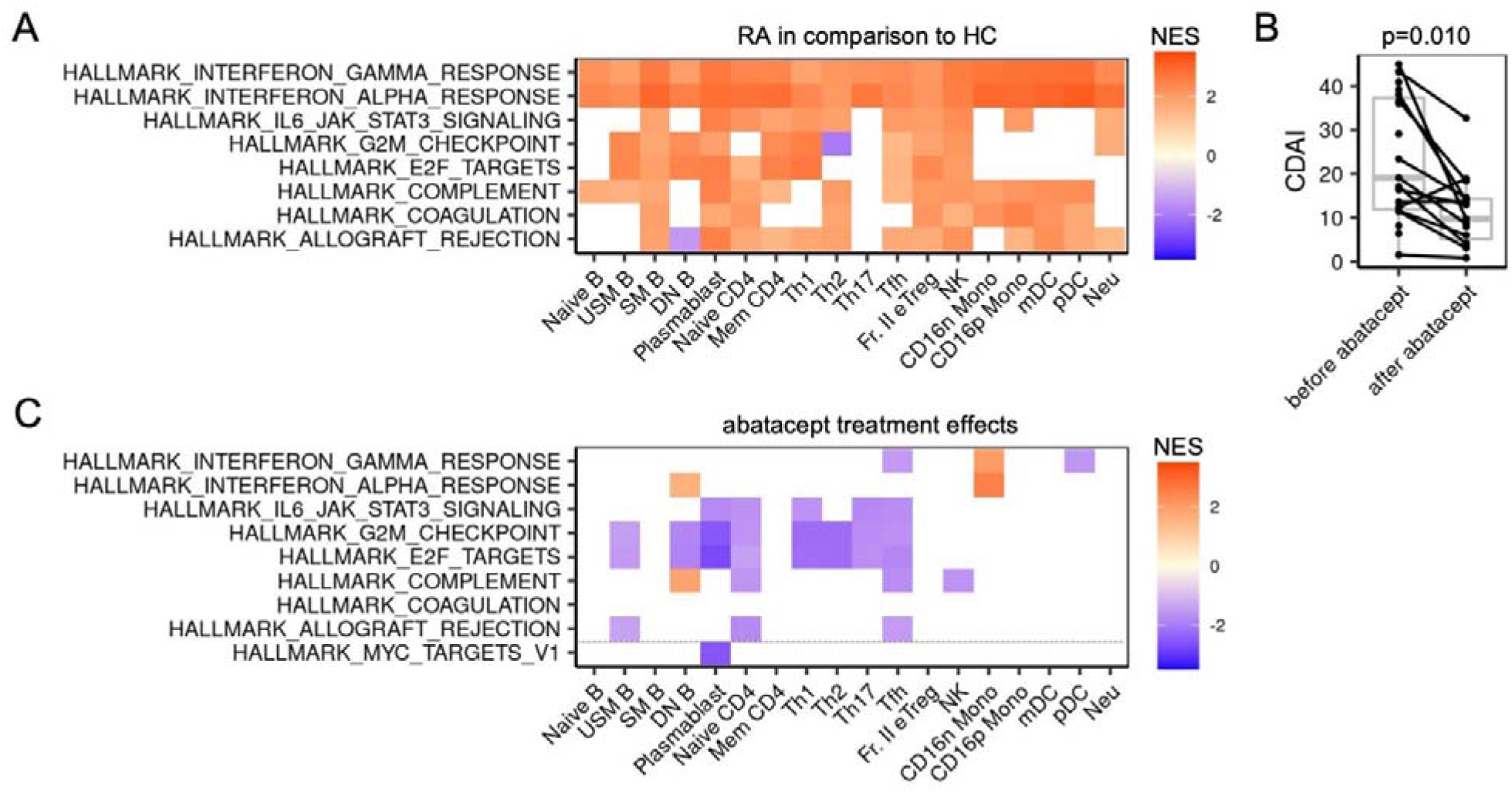
RA immune cell gene expression and abatacept treatment-induced partial normalization. (A) The GSEA results in the RA population prior to treatment compared to the HC population for each subset. Gene sets with |NES| > 2.5 in at least 1 subset were targeted; white indicates that enrichment was not significant. (B) Clinical treatment effects of ABT. (C) The GSEA results for RA before and after treatment with ABT. The 8 gene sets from (A) with increased expression in the RA population and the gene sets with a change in the |NES| > 2.5 in at least 1 subset are shown. Pathways with FDR < 0.05 are colored. RA: rheumatoid arthritis, HC: healthy control, NES: normalized enrichment score, GSEA: gene set enrichment analysis, ABT: abatacept. Definitions of the subsets are provided in online supplementary table 2.

Next, we performed GSEA on the immune cells before and after treatment with ABT (n = 15, figure 2B and 2C). Notably, although ABT treatment significantly improved disease activity, ABT did not suppress the expression of IFN-response genes before and after treatment (figure 2C). ABT tended to decrease the expression of genes related to various inflammatory responses including IL6-JAK-STAT3 signaling and cell proliferation in B and CD4^+^ T cell subsets (figure 2C). A decrease in expression of MYC-associated genes were seen in plasmablasts after treatment.

In TCZ treated RA patients, inflammatory response genes were down-regulated in monocytes, although the result was statistically limited by the small number of patients in the TCZ treatment group (n=5, online supplementary figure 2).

### Pre-DC genes in peripheral blood are associated with poor treatment prognoses

To clarify baseline characteristics of the immune cells in RA with a poor treatment response, we defined patients who had achieved CDAI50 as responders (24, 25). No significant background clinical differences existed between the responders and the non-responders (online supplementary table 3). We prepared co-expression gene modules based on a weighted gene co-expression network analysis (WGCNA) for each of the immune cell subsets in the RA patients, and investigated the associations with future treatment response.

564 modules were constructed with WGCNA, and the strongest association with treatment-resistance was identified for a module “pDC_M18” (figure 3A). The pDC_M18 genes were expressed at higher levels in treatment-resistant RA compared to HC (figure 3B). Notably, treatment did not significantly affect pDC_M18 score (figure 3C). Additionally, baseline pDC_M18 was a better predictor of response than anti-CCP antibody (anti-citrullinated protein antibody [ACPA]) or disease duration, which are established clinical parameters for resistance to therapy (figure 3D) (26-28). In addition, we found no significant association between pDC_M18 expression and any clinical measures (online supplementary figures 3A to 3F), although the pDC_M18 expression tended to be higher in patients with longer disease duration than in patients with shorter disease duration (with the cutoff being defined as 1 year from onset) (29) (*p* = 0.068) (online supplementary figure 3F). Moreover, pDC_M18 tended to predict treatment resistance regardless of the type of molecular targeted drug (online supplementary figure 3G). The pDC_M18 module consisted of 207 genes (online supplementary table 4) and the network of the top 50 hub genes is shown in figure 3E. Pre-DC is a subset that shares many of the same markers as pDC and differentiates into cDC1 and cDC2 (20). Unexpectedly, the hub genes of pDC_M18 included a number of pre-DC signature genes, such as *CD33, CX3CR1, KLF4*, and *CD22*. In fact, 13 of the genes were included in the pre-DC signature genes reported by See P *et al*. (20) (online supplementary table 5) (*CD22, CD244, CD33, CD63, CD93, CLEC10A, CLEC12A, CX3CR1, ITGAX, KLF4, KLF8, RAB32*, and *SIGLEC6*; odds ratio [OR] = 48.8; *p* < 2.2e-16). Of the pDC WGCNA modules, only pDC_M18 exhibited significant overlap with the pre-DC signature genes (OR = 27.4, *p* = 1.0e-13; figure 3F). In addition, a clear correlation was found with the expression level of ME of this module for the proportion of pre-DC in the pDC sample, estimated by deconvolution applying the data from the paper of See P *et al*. using CIBERSORTx (30) (r = 0.70, *p* = 9.0e-8) (figure 3G). Pre-DC is contained in the subset that have conventionally been considered pDC (20). It was therefore thought that pDC_M18 reflects the proportion of pre-DC in pDC.

**Figure 3.**
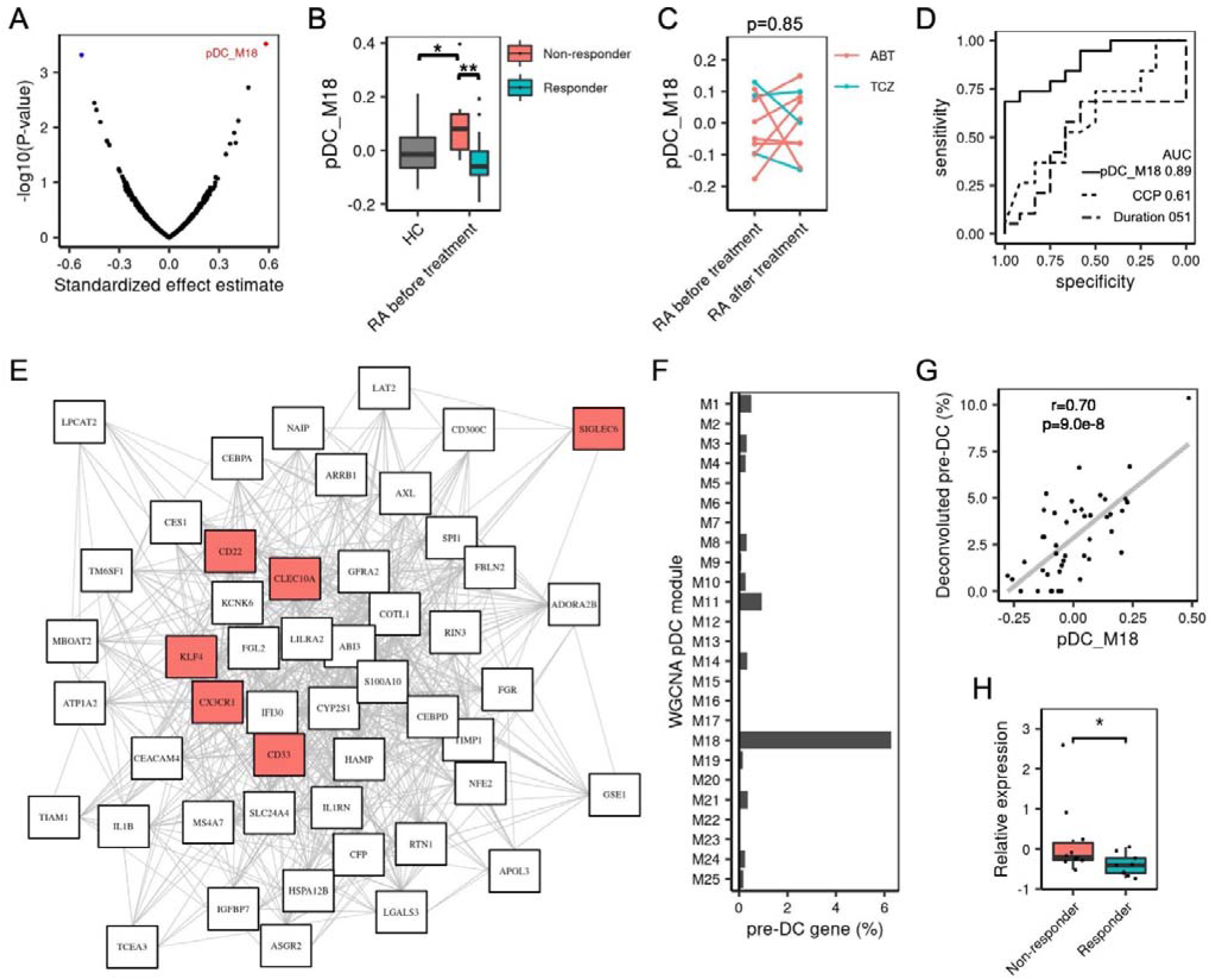
Pre-DC genes correlate to RA treatment resistance. (A) The assessment was performed using a generalized linear model with failure to achieve CDAI50 at 6 months as the target variable and the eigengene of the module of each immune cell subset as the explanatory variable. Color-coding was performed with Benjamini-Hochberg FDR < 0.10 as the significance level. (B) Comparison of ME expression in the pDC_M18 module in the pre-treatment RA populations and the HC population. (C) The change over time in pDC_M18 expression associated with treatment was evaluated in 3 patients who received TCZ and 7 patients who received ABT from whom blood samples were obtained before and after treatment. Paired t-test. (D) ROC curve of pre-treatment pDC_M18 expression and treatment prognosis. The dotted line is that of the anti-CCP antibody positive and treatment prognosis, and the dotted and dashed line is that of longer disease durations (≥ 1 year) and treatment prognosis. (E) Network figure of gene expression correlations for the top 50 hub genes in the pDC_M18 module. The pre-DC signature genes are color-coded. Gene pairs with a Pearson’s correlation coefficient of expression > 0.6 were connected with each other. (F) Match rate with pDC_M18 genes in each pDC WGCNA module. (G) Correlation of the pDC_M18 ME and the proportion of deconvoluted pre-DC in the paper of See P et al. (20). (H) qPCR was performed on the pDC in peripheral blood before treatment in a separate confirmatory cohort (n = 19) of RA patients before starting a new therapy, and the pDC_M18 expression signatures of the non-responder and responder groups were compared. *p < 0.05, **p < 0.01. pDC: plasmacytoid dendritic cell, HC: healthy control, RA: rheumatoid arthritis, ABT: abatacept, TCZ: tocilizumab, AUC: area under the curve, WGCNA: weighted correlation network analysis, ME: module eigengene.

AS DC is a subfraction of DC that overlaps with pre-DC (22) and is characterized by the expression of *AXL, SIGLEC1*, and *SIGLEC6* and powerfully activates T cells (21). The signature genes of AS DC, defined by Villani AC *et al*., are enriched in the pDC_M18 module with 13 overlapping genes (OR = 12.5; *p* = 4.4e-10; *ACPP, ADAM33, AXL, CD22, CX3CR1, CXCR2, FAM129A, GPR146, HIP1, KLF4, S100A10, SIGLEC1*, and *SIGLEC6*) (online supplementary table 5, online supplementary figure 4A). The pDC_M18 module can therefore reflect the proportion of AS DC, similar to that of pre-DC (r = 0.69, *p* = 1.3e-7) (online supplementary figure 4B).

**Figure 4.**
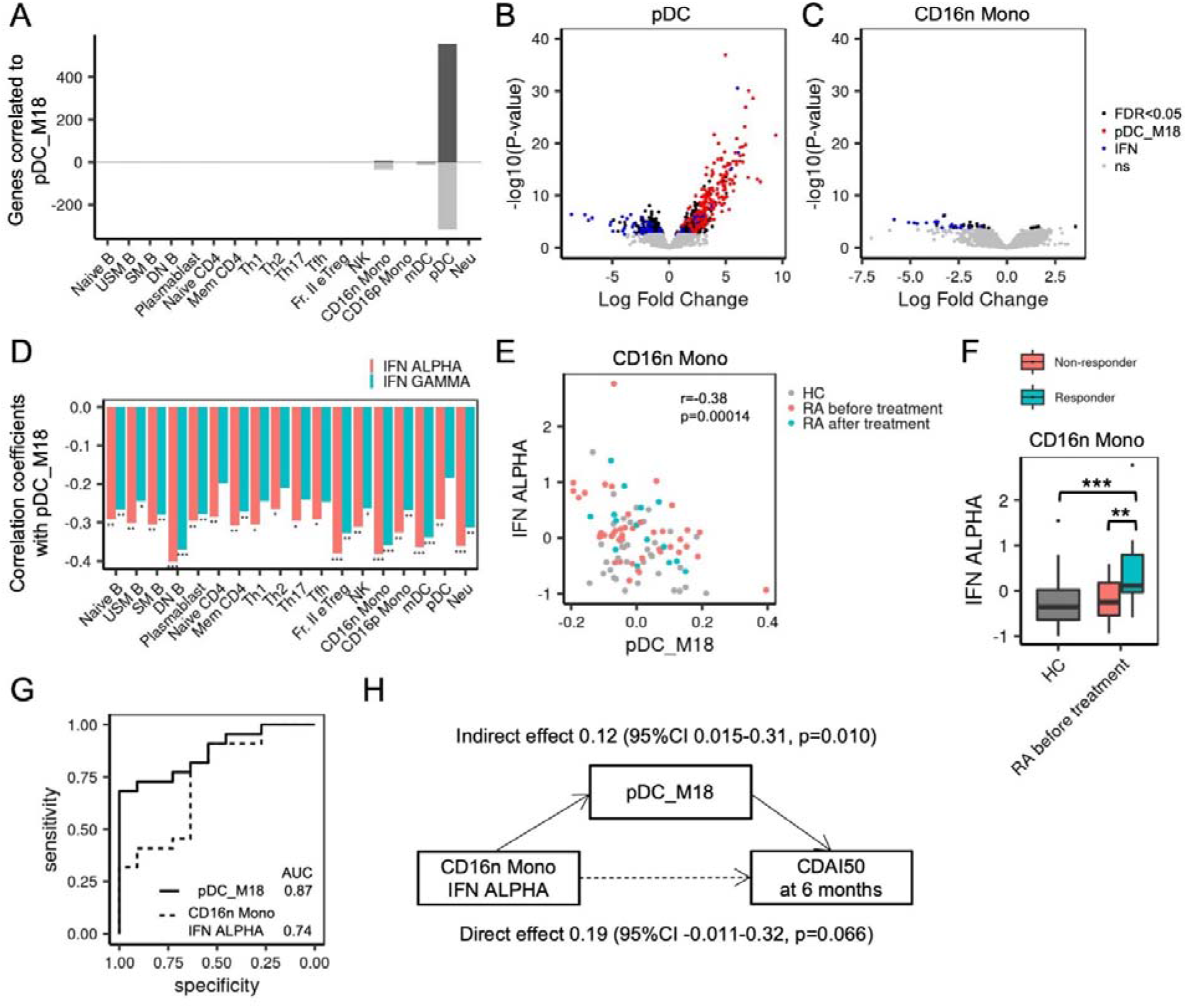
Inverse correlation of pDC_M18 and IFN response genes. (A) Number of genes for which there was a relationship between the pDC_M18 and gene expression. The number of genes for which there was a positive correlation to pDC_M18 (positive values, shown in dark grey) and the number of genes for which there was a negative correlation to pDC_M18 (negative values, shown in light grey) are shown separately. (B-C) pDC, CD16n Mono volcano plot, showing gene expression correlated to pDC_M18. The genes in pDC_M18 (red) and the genes associated with pDC_M18 (FDR with Benjamini-Hochberg < 0.05) that were IFN response genes (blue) were color-coded separately. (D) Correlation coefficients for the IFN response signature and pDC_M18 for each immune subset. (E) Negative correlation between the IFN-α response signature and pDC_M18 in CD16n Mono. (F) Comparison of the IFN-α response signature in CD16n Mono in the pre-treatment RA populations and the HC population. (G) ROC curve of pre-treatment pDC_M18 and CD16n Mono IFN-α response signature expression and treatment prognosis. (H) Mediation model representing the relationships between CD16n Mono IFN-α response signature expression, pDC_M18, and CDAI50 at 6 months. *p < 0.05, ** p < 0.01, *** p < 0.001. IFN: interferon. Subset definitions are provided in online supplementary table 2.

In other words, the cells fractionated as pDC contain cell fractions referred to as pre-DC and AS DC. The fact that these cells are increased in treatment-resistant RA patients is reflected in the gene expression profile.

### Confirmation of the pDC treatment-resistant module with an independent cohort

Next, in a validation cohort (n = 19) of RA patients prior to the initiation of new treatment, we performed quantitative polymerase chain reaction (qPCR) for the pDC in peripheral blood collected before treatment and evaluated the mean Z scores of the expression of the hub genes in pDC_M18. No significant differences in pre-treatment activity were found between the responders (n = 9) and the non-responders (n = 10) (online supplementary table 6). The pDC_M18 hub genes were expressed in significantly higher levels in the treatment-resistant group (*p* = 0.043, figure 3H), and the relationship between high expression of pre-DC genes in the pDC fraction and treatment resistance was reproduced.

### Inverse correlation between pre-DC gene expression and IFN gene expression

To investigate the effects of pDC_M18 expression on immune cells, we searched for genes associated with pDC_M18 expression in each immune cell subset in the study cohort. A high level of pDC_M18 expression inversely correlated to a low level of expression of IFN-related genes, particularly in pDC and CD16n Mono (figures 4A to 4C, and online supplementary figure 5). Additionally, in all immune cell subsets studied, an inverse correlation was found between genes included in the pDC_M18 module and the IFN-α response signature (figures 4D and 4E). The IFN-α response signature expression was increased in responder RA patients, particularly in CD16n Mono (figure 4F, online supplementary figure 6). Although type I IFN gene signature has been proposed as predictive factor for treatment response (5, 9, 10), IFN-α response signature expression in CD16n Mono only showed modest predictive power for CDAI50 at 6 months (figure 4G).

Using mediation analysis, we tested whether IFN-α response signature of CD16n Mono has a direct effect on CDAI50 response at 6 months or was indirectly mediated by pDC_M18. pDC_M18 significantly mediated the effects of IFN-α response signature expression on CDAI50 (*p* = 0.010) (figure 4H).

### An increase in the pre-DC cell population in peripheral blood predicts treatment resistance

Next, we used mass cytometric analysis to further validate the relationship between the populations of immune cells in peripheral blood prior to treatment and treatment prognosis in a second validation cohort (n = 28) of RA patients who were going to start ABT therapy. When we compared the clinical characteristics from the responders (n = 21) to those from the non-responders (n = 7) (online supplementary table 7), no significant differences were found between the two groups in clinical findings except for age, which was lower in the treatment-resistant group (*p* = 0.017).

The cells were clustered based on data on the expression of 36 cell surface proteins obtained through mass cytometry, and the immune cells in the peripheral blood were classified into 27 populations (figures 5A and 5B, and online supplementary figure 7). However, no significant relationships to treatment prognosis were found for any of these populations (figure 5C).

**Figure 5.**
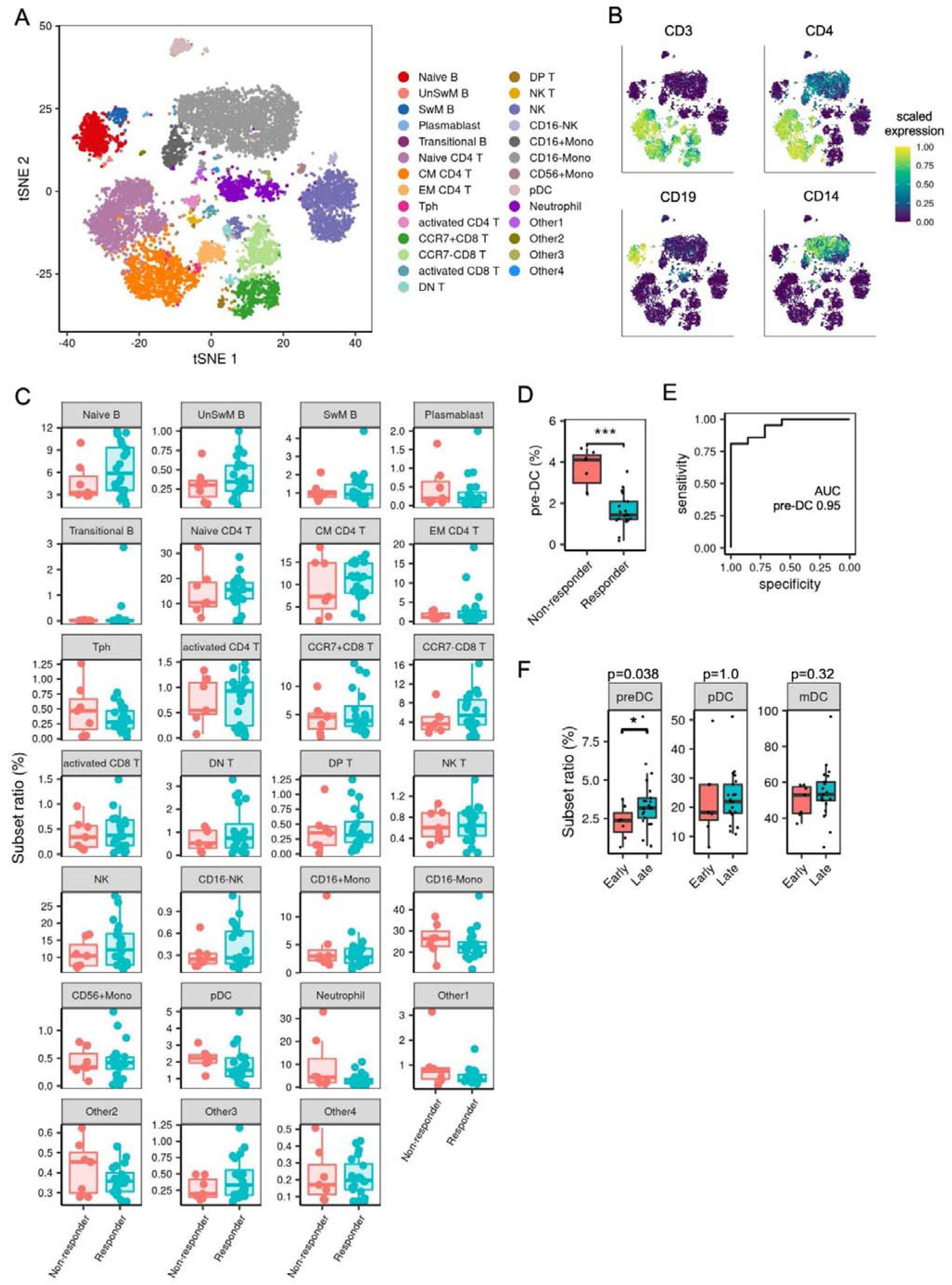
Search of immune cells associated with prognosis by mass cytometry of RA patients before ABT treatment initiation. (A) tSNE plot of the peripheral blood immune cell population in 28 RA patients before ABT therapy. The mass cytometry data for 36 cell surface markers were clustered, and 27 cell populations identified. (B) Representative plots of several cell surface markers. The same tSNE plots as those described in (A) were used. (C) Comparison of the proportion of the 27 cell populations with treatment prognosis (achievement of CDAI50 after 6 months). (D) Comparison of proportions of pre-DC relative to DC with treatment prognosis. (E) ROC curve for the proportion of pre-DC before treatment and treatment prognosis. (F) The proportions of pre-DC, pDC, and mDC in the longer (> 1 year) or shorter (≤ 1 year) disease duration groups were compared. tSNE: t-distribution Stochastic Neighbor Embedding.

Because we were not able to identify pre-DC in the automated clustering analysis, we performed manual gating, defining Lin (CD34, CD3, CD14, CD19, CD16)^−^ HLA-DR^+^ CD45RA^+^ CD123^+^ CD11c^nega/mid^ CX3CR1^+^ as pre-DC (online supplementary figure 8), which was consistent with the pre-DC gating used in the paper of See P *et al*. (20) (online supplementary figure 9).

The proportion of pre-DC relative to DC in the peripheral blood obtained prior to treatment was significantly higher in the treatment-resistant group (*p* = 4.0e-06) (figure 5D and online supplementary table 8). The proportion of pre-DC prior to treatment also had superior prognosis predictive performance with an area under the curve (AUC) of 0.95 (figure 5E). Additionally, the proportion of pre-DC was higher in patients with a longer disease duration (*p* = 0.038) (figure 5F).

### Synovial pre-DC-like cells are inflammatory cDC2s

Finally, to investigate the pathophysiological relevance of pre-DC in arthritis, we analyzed previously reported synovial single cell RNA-seq data of treatment naïve RA patients (n=16) (31). We separated DCs from CD45^+^ synovial immune cells, and clustering analysis identified 10 DC clusters (online supplementary method, figure 6A and 6B). The *IL3RA* expression clearly distinguished pDC from cDCs. A cluster of cDCs characteristically expressed pDC_M18 genes, such as *CLEC12A* and *CD33*, and we named them as “pre-DC-like” cells (figure 6B and 6C). Cluster marker genes, such as *CLEC9A* for cDC1(32), clearly distinguished cDC1 clusters (CLEC9A^+^ DC and S1008B^+^ DC) from cDC2 clusters (pre-DC-like, SPP1^+^ DC, CXCL8^+^ DC, and cDC) (figure 6B and figure 6D). LAMP3^+^ DCs uniquely expressed *LAMP3*, a reported marker of regulatory DC that limit antitumor immunity (33). Recently, cDC2 are subdivided to two clusters; anti-inflammatory cDC2A and pro-inflammatory cDC2B (34). Synovial pre-DC-like cells and other cDC2 populations expressed signature genes of proinflammatory cDC2B (figure 6E). On average, 32 percent of the Synovial DCs were pre-DC-like, and the frequency was higher in RA patients in longer duration (with the cutoff being defined as 3 years from onset here) (figure 6F, *p* = 0.16), consistent with our peripheral blood data.

**Figure 6.**
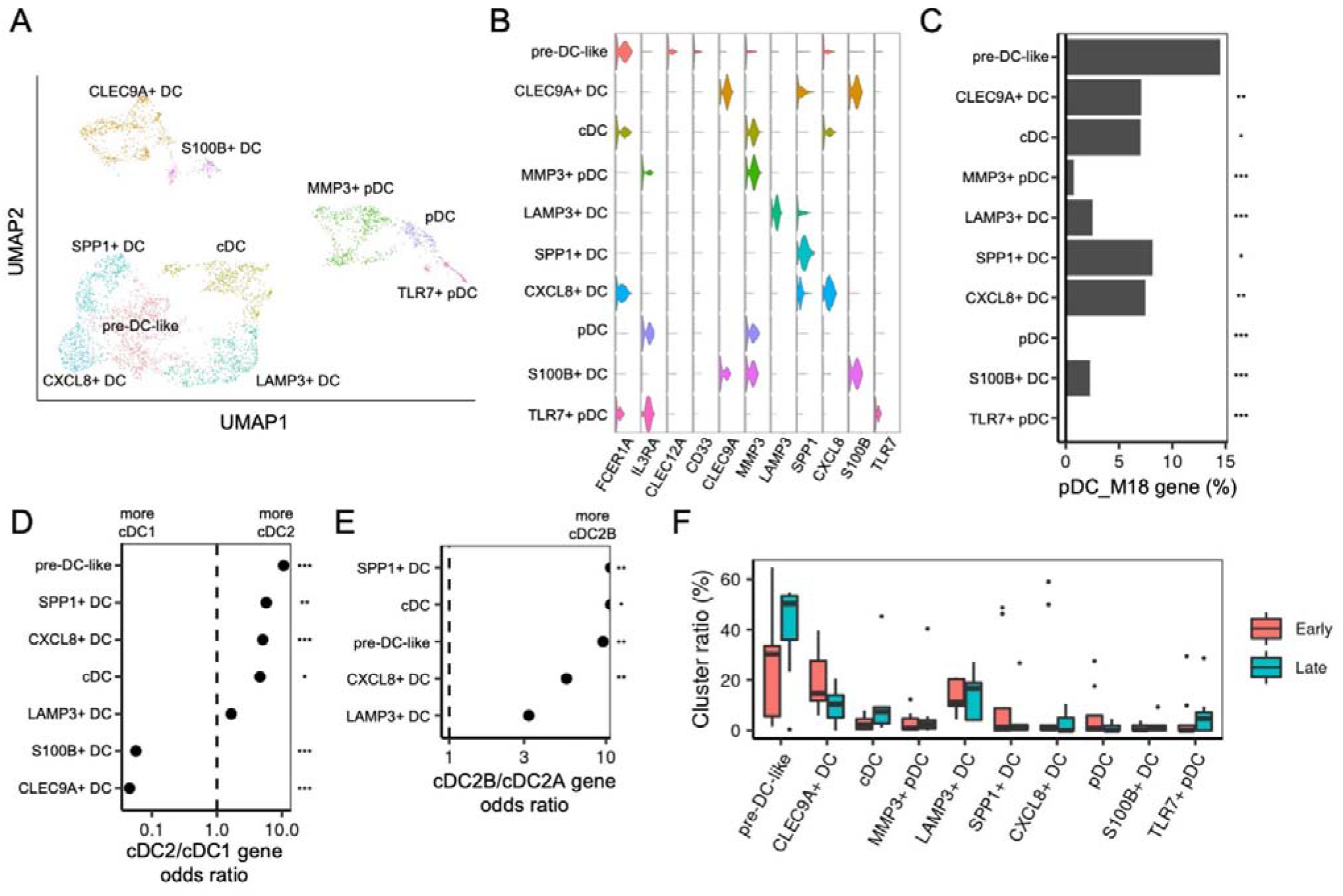
Synovial dendritic cells single cell RNA-seq analysis of untreated RA. (A) UMAP plot of synovial DCs from untreated rheumatoid arthritis patient (n=16, 3804 cells). (B) Violin plots of DC cluster marker gene expressions. (C) Match rate with pDC_M18 genes in each DC cluster signature genes. The match rate of pre-DC-like cluster was compared with other DC clusters with Fisher’s exact tests. (D-E) Odds ratios of cDC2 and cDC1 signature genes (D) or pro-inflammatory cDC2B and anti-inflammatory cDC2A signature genes (E) in each DC cluster signature genes. (F) DC cluster proportions, stratified by disease duration (duration < 3 years = Early, n=9). *p < 0.05, ** p < 0.01, *** p < 0.001.

## Discussion

In this study, we characterized the gene profiles of the immune cells of RA patients and demonstrated that treatment resistance can be predicted by an increase in pre-DC in the peripheral blood of RA patients prior to starting therapy. This result shows the potential for realizing a stratified therapy of RA based on analysis of pre-DC in peripheral blood, which is minimally invasive, and also shows that pre-DC may be involved in the immunopathology of treatment-resistant RA.

No significant differences were found between the poor treatment prognosis group and the good treatment prognosis group in rheumatoid factor (RF), ACPA or disease duration (online supplementary table 3), and pre-DC was superior for predicting the prognosis than these known clinical prognosis predictive factors (figure 3D). Although ACPA and RF positivity are established as poor prognosis factors (26, 27, 35), pre-DC may be associated with treatment resistance via a distinct mechanism. Moreover, the results of this study suggest that there is an increase in pre-DC in the peripheral blood prior to treatment in patients with a long (≥ 1 year) disease duration. This is consistent with reports that the duration of disease tends to be longer in treatment-resistant patients (28). It is known that in early RA there is a so-called window of opportunity in which treatment response is good (36). An increase in pre-DC may have an immunological foundation, where treatment response worsens as the disease duration increases. Delay to initial treatment is a risk factor of refractory RA (3), which can also be associated with pre-DC.

Recently, the human DC have been categorized in greater detail. Villani AC *et al*. used scRNA-seq to establish 6 populations (21). AS DC, a population that is comparable to pre-DC, has been shown to be increased in pediatric systemic lupus erythematosus (SLE) patients (37). Pro-inflammatory cDC2s, that can present antigen to CD4 T cells and are known both as cDC2B or cDC3 (32), were expanded in SLE and were correlated with disease activity (38). In our synovial single-cell transcriptome profiling, we identified pre-DC-like DCs occupied around a third of synovial DC population and that were related to pro-inflammatory cDC2Bs (figure 6). cDC2Bs are suggested to mediate Th17 responses (34). In treatment-resistant patients, an increase in pre-DC may affect the synovial acquired immune response in RA by stimulating the CD4^+^ T cells, either as pre-DC-like cells themselves, or after further differentiating into pro-inflammatory cDC2s. It has been reported that, in a mouse model of influenza A virus infection, pre-DC enter infected tissues (39), and it is possible that, in human arthritis, as well, there is a link between an increase in pre-DC, not only in the blood but also at the joint, and inflammation.

In this study, we observed a negative correlation between type I IFN signaling genes and pre-DC gene expression, and pre-DC gene expression had a closer relationship with treatment response than IFN-α signaling (figure 4). It is possible that IFN signaling is inhibitory on pre-DC, or that pre-DC are antagonistic to the pDC that produces IFN-α. The type I IFN stimulation is reported to limit cDC differentiation and promotes pDC differentiation (40) (41). Although type I IFN signaling has been reported to be a factor that predicts development of RA (42), type I IFN signaling predicts a good therapeutic response to TCZ and TNF inhibitors (5, 9, 10). The extent of type I IFN signaling moreover differs considerably depending on the individual RA patient (23), and it has been reported that around 33% of RA patients exhibit increased type I IFN signaling (43). It is possible that there are separate RA subtypes with different immunolopathologies: RA with a relatively good prognosis characterized by increased type I IFN signaling; and RA with a relatively poor prognosis characterized by an increase in pre-DC.

One limitation of this study was that the patients’ existing treatments and the new treatments they were going to start receiving were not the same, and the analyses were therefore performed using combined responses to various treatments. In the future, a larger-scale investigation that also takes into account differences in patients’ clinical pictures and/or existing treatments should be conducted.

In conclusion, we discovered, through an analysis of the gene expression profiles of immune cells in the peripheral blood of patients prior to treatment, that an increase in pre-DC can predict the prognosis of RA, and the reproducibility of this result was confirmed using two cohorts. We also identified synovial pre-DC-like cells that were similar to proinflammatory cDC2 in transcriptome. These results can contribute to our understanding of RA treatment stratification and the pathology of difficult-to-treat RA.

## Supporting information

supplementary_manuscripts

## Data Availability

The datasets generated during this study are available at the National Bioscience Database Center (NBDC) with the study accession code of hum0214. We used publicly available software for the analyses.

## Acknowledgements

We thank generous supports for sample and clinical data collection from Dr. H Shirai, Dr. J Maeda, Dr. M Ogawa, Dr. Haruka Takahashi, Dr. A Nishiwaki, Dr. Hideyuki Takahashi, Dr. T Itamiya, Dr. Y Akutsu, and Ms. H Ikeda of the University of Tokyo, and Mr. T. Ura of Azuma Rheumatology Clinic. The super-computing resource was provided by Human Genome Center, Institute of Medical Sciences, The University of Tokyo (http://sc.hgc.jp/shirokane.html).

## Author Contributions

YN and KF designed the study. SY and YN conducted bioinformatics analysis with the help of M Ota, HH. SY, YN, M Ota, HH, Y Takeshima, M Okubo, SK, YS, MN, RY, NH, YI, SS, and HK contributed to sample and clinical data collection for RNA-seq analysis. Y Tsuchida, YS, and KI contributed to critical reading and revision of the manuscript. KK, K Shimane, K Setoguchi and TA contributed sample and clinical data collection for mass cytometry analysis. YN, MW, and XZ contributed to synovial single cell RNA-seq data analysis. HS, KY, TO, and KF managed the project. SY and YN wrote the first manuscript with critical inputs from M Ota, TO and KF. All authors contributed to the final version of the manuscript and approved it.

