## supplementary_manuscripts for "Immunomics analysis of rheumatoid arthritis identified precursor dendritic cells as a key cell subset of treatment resistance"

**Online supplementary materials and methods**

**Study cohorts**

The study population consisted of adult healthy control (HC) volunteers and adult rheumatoid arthritis (RA) patients being treated on an outpatient basis by the Allergy and Rheumatology Department of the University of Tokyo Hospital. This study was a prospective study that enrolled RA patients who met the 2010 rheumatoid arthritis classification criteria of the American College of Rheumatology (ACR)/European Alliance of Associations for Rheumatology (EULAR) (1). Peripheral blood samples were collected from HC (n=39) and RA subjects (n=55) before they started receiving new therapies. Of those, 22 received abatacept (ABT), 12 received tocilizumab (TCZ), 7 received tumor necrosis factor (TNF) inhibitors, 6 received tofacitinib, 6 received methotrexate, and 2 received other conventional synthetic disease-modifying antirheumatic drugs (csDMARDs). Peripheral blood samples were collected from 20 of the RA subjects 6 months after they have started receiving new therapies: 15 patients on ABT and 5 patients on TCZ.

Although there are various measures of RA disease activity, the CDAI does not incorporate C-reactive protein (CRP), and therefore is not influenced by infections and/or the use of interleukin (IL)-6 inhibitors. In this study, therefore, we defined subjects who had achieved improvements of 50% or more on the CDAI after 6 months of treatment as responders (treatment-responsive patients), and patients who had achieved improvements of less than 50% on the CDAI as non-responders (treatment-resistant patients) (2, 3). Subjects who had been treated with biological DMARDs, targeted synthetic DMARDs, or methotrexate (MTX) were included in the analysis of treatment resistance; 1 subject who had been treated with iguratimod (IGU), 1 subject who had been treated with salazosulfapyridine (SASP), and subjects for whom no data were available on their clinical prognoses at 6 months were excluded from the analysis.

Persons were eligible to participate in the study as HC if they had no past treatment history and were not taking any medications, including supplements.

This study was approved by the Ethics Committees of the University of Tokyo (G-10095, G-10084). Written informed consent was obtained from each subject in accordance with Declaration of Helsinki.

**Patient and Public Involvement Statement**

Patients or the public were not involved in this study.

**PBMC collection and RNA purification**

Whole blood in an amount of 30 mL was mixed with 1 mL of heparin and diluted with an equivalent amount of wash buffer (phosphate-buffered saline + 2% fetal calf serum [BioWest] + 1 mM EDTA [ethylenediaminetetraacetic acid]), and the peripheral blood mononuclear cell (PBMC) layer was recovered by density gradient centrifugation (1000 g for 10 minutes at 20$℃$) using the Ficoll-Plaque PLUS (GE Healthcare). Following hemolysis treatment with an ammonium chloride potassium solution (150 mL NH_4_Cl + 10 mM KHCO_3_ + 0.1 mM Na_2_EDTA), the resulting product was washed using wash buffer. Following non-specific binding inhibition of Fcγ receptors by means of a human Fc receptor binding inhibitor (eBioscience), the resulting product was stained using various fluorescently labelled antibodies. After staining, the resulting product was suspended in a basic sort buffer (Hank’s balanced salt solution + 2% fetal calf serum + 1 mM EDTA + 25 mM 4-(2-hydroxyethyl)-1-piperazineethanesulfonic acid), and flow cytometry was used for analysis and cell sorting.

**Sorting of immunocompetent cell subsets by flow cytometry**

The data that were included in this analysis consisted of 4 datasets obtained using different flow cytometry and sorting procedures, which were defined as Datasets 1 to 4 (online supplementary table 9).

Datasets 1 and 2 consisted of data newly obtained for this study, and Datasets 3 and 4 consisted of data that had already been published as part of ImmuNexUT (4).

In Datasets 1 to 3, the MoFlo XDP sorter (Beckman Coulter) was used to sort the following 19 types of cell subsets: Naïve CD4 T cells (Naïve CD4), Memory CD4 T cells (Mem CD4), T helper 1 cells (Th1), T helper 2 cells (Th2), T helper 17 cells (Th17), T follicular helper cells (Tfh), Fraction II effector regulatory T cells (Fr. II eTreg), Naïve CD8 T cells (Naïve CD8), Memory CD8 T cells (Mem CD8), Naïve B cells (Naïve B), Unswitched memory B cells (USM B), Switched memory B cells (SM B), Double negative B cells (DN B), Plasmablasts (Plasmablast), Natural killer cells (NK), CD16 positive monocytes (CD16p Mono), CD16 negative monocytes (CD16n Mono), Myeloid dendritic cells (mDC), and Plasmacytoid dendritic cells (pDC). Definitions of each of these cell subsets, based on cell surface antigens, are provided in online supplementary table 10.

Dataset 4 consisted of 26 cell subsets, sorted using the FACS Aria^TM^ Fusion flow cytometer: Naïve CD4 T cells (Naïve CD4), Memory CD4 T cells (Mem CD4), T helper 1 cells (Th1), T helper 2 cells (Th2), T helper 17 cells (Th17), T follicular helper cells (Tfh), Fraction II effector regulatory T cells (Fr. II eTreg), Fraction I naïve regulatory T cells (Fr. I nTreg), Fraction III non-regulatory T cells (Fr. III T), Naïve CD8 T cells (Naïve CD8), CD8+ T effector memory CD45RA+ cells (TEMRA CD8), Effector memory CD8 T cells (EM CD8), Central memory CD8 T cells (CM CD8), Naïve B cells (Naïve B), Unswitched memory B cells (USM B), Switched memory B cells (SM B), Double negative B cells (DN B), Plasmablasts (Plasmablast), Natural killer cells (NK), CD16 positive monocytes (CD16p Mono), Non-classical monocytes (NC Mono), Intermediate monocytes (Int Mono), CD16 negative monocytes (CD16n Mono), Myeloid dendritic cells (mDC), Plasmacytoid dendritic cells (pDC), and Low-density granulocytes (LDG). Definitions of each of these cell subsets, based on cell surface antigens, are provided in online supplementary table 11. Each subset was sorted with the upper limit being 5,000 cells, and then lysed and stored in a deep freezer at -80$℃$. Furthermore, the fluorescently labeled antibodies that were used in PBMC staining for each dataset are shown in online supplementary table 12.

When the data from Datasets 1 to 3 were combined with the data from Dataset 4, the subsets that had been collected only in Dataset 4 (Fr. I nTreg, Fr. III T, TEMRA CD8, EM CD8, CM CD8, NC Mono, Int Mono) were excluded from the analysis. In addition, because the definitions of the Naïve CD8 and Mem CD8 subsets in Datasets 1 to 3 were different from the definitions in Dataset 4, the subsets collected in Datasets 1 to 3 were excluded from the analysis. The 18 subsets that were ultimately analyzed are shown in online supplementary table 2.

**Collection of peripheral blood neutrophils**

Samples of 3 mL of whole blood were collected using collection tubes containing EDTA-2K. For Dataset 1, the samples were subjected to magnetic separation using EasySep direct human neutrophil isolation kits (STEMCELL Technologies). For Datasets 2 to 4, the samples were subjects to magnetic separation using “MACSexpress Neutrophil Isolation Kit, human” kits (Miltenyi Biotec), and the erythrocytes were removed using “MACSexpress Erythrocyte Depletion Kit, human” kits (Miltenyi Biotec). From each sample, around 2×10^6^ cells were then obtained, and lysed using TRIZOL LS reagent (Invitrogen), and the resulting product was stored in a deep freezer at -80$℃$.

**cDNA library preparation and RNA-sequencing (RNA-seq)**

Each recovered cell subset sample was subjected to RNA purification using NucleoSpin RNA (Takara Bio) (Dataset 1), RNeasy Micro Kit (QIAGEN) (Datasets 2 and 3), or MagMAXTM96 Total RNA Isolation Kit (Thermo Fisher Scientific) (Dataset 4).

Sequence libraries were prepared using the SMART-seq V4 Ultra Low Input RNA Kit for Sequencing (Clontech Laboratories, Inc.), the Nextera XT DNA Library Prep Kit (Illumina, Inc.), or the Nextera XT Index Kit V2 Set A-D (Illumina, Inc.). The HiSeq 2500 (Illumina, Inc.) (Datasets 1, 3, and 4) and the NovaSeq 6000 (Illumina, Inc.) (Dataset 2) systems were used to obtain 100 bp paired-end base sequences. The bcl files were demultiplexed and converted into FASTQ files using bcl2fastq2 conversion software v2.17.

**FASTQ file quality control and mapping and counting**

The adapter sequences were removed using Cutadapt v1.14 (5), and FASTX Toolkit v0.0.14 was used to remove bases with a Phred quality score below 20 from the 3’-terminal (http://hannonlab.cshl.edu/fastx_toolkit/), and reads with a base length of less than 50 base sequences were removed. Reads having 20% or more bases with a Phred quality score below 20 were also removed. The USCS human genome 38 (hg38) was used as the reference sequence, mapping was performed using STAR v2.5.3a (6), and counting was performed using HTSeq v0.9.1 (7). Samples with a uniquely mapped rate below 80% or fewer than 5,000,000 uniquely mapped reads were excluded from subsequent analyses.

**Quality control and normalization of RNA-seq samples**

Low expressing genes having counts below 10 in 10% or more of samples were excluded from analysis. In addition, within each subset, the mean correlation coefficient (D score) between each sample and all of the other samples was calculated, and samples with a D score that was below the mean – 2sd of all of the samples were excluded from analysis as outliers. The average D score cutoff value was 0.90. The gene expression level was normalized by Trimmed mean of M values (TMM) (8), the counts were converted to log2-transformed counts per million, dataset-dependent batch effect correction was performed for subsets other than Neu, and reagent-dependent batch effect correction was performed for Neu (Dataset 1 vs. Datasets 2 to 4) using the sva package comBat function (9). High expression levels of the characteristically expressed genes were consistent with definitions of each of the cells (online supplementary figure 1A). Principal component analysis (PCA), a dimensional compression technique, was performed on each subset, and 1 clearly outlying sample was excluded from Neu. A total of 1701 samples (622 from HC, 797 from RA patients before treatment, and 282 at 6 months after treatment) were included in the analysis (online supplementary figure 1B and online supplementary table 13). For all of the samples, PCA was performed using the most highly variable 500 genes.

**RNA-seq sample variance decomposition**

Variance decomposition of the normalized data was performed using variancePartition v1.12.3 (10). The following linear mixed model was used to calculate the fixed effect of age and the random effects of subset, individual, disease (the difference between RA and HC), sex, and batch (Dataset 1 to 4) on gene expression.

Gene expression ~ (1|subset) + (1|individual) + (1|disease) + age + (1|sex) + (1|batch)

**RNA-seq sample differentially expressed gene set analysis**

The RA before treatment population was compared to the HC population first, and the RA after ABT treatment population was then compared to the RA before ABT treatment population (n=15). In the same way, RA population before and after TCZ populations were compared (n=5). A differentially expressed genes (DEGs) analysis was performed for each subset using edgeR v3.24.3, and the log fold change of each gene was calculated (8). Additionally, for each subset, a Gene Set Enrichment Analysis (GSEA) was performed for the MSigDB hallmark gene set (msigdbr v7.4.1) (11, 12).

Normalized gene expression Z scores were prepared for the gene sets contained in MSigDB HALLMARK_INTERFERON_ALPHA_RESPONSE and HALLMARK_INTERFERON_GAMMA_RESPONSE, HALLMARK_IL6_JAK_STAT3_SIGNALING, and the means thereof were considered the gene set signatures.

**Gene module analysis by weighted correlation network analysis (WGCNA)**

A weighted correlation network analysis (WGCNA) was performed using version 1.68 and the RNA-seq data from the RA before treatment population to explore what gene modules are associated with treatment resistance (13). For the CD4 T cell subsets Th1, Th2, Th17, and Tfh, because the number of samples from the pre-treatment RA population was limited (< 30) (online supplementary table 13), and because these subsets were similar to the Mem CD4 subset in terms of gene expression (figure 1B), they were excluded from this analysis. For each cell subset, the groups of genes for which the changes in expression were positively correlated were considered modules, and module eigengene (ME) was used as the representative value for the expression of each module. A generalized linear model with failure to achieve CDAI50 at 6 months after treatment initiation as the target variable and each immune cell subset ME as the explanatory variable were used to calculate the standardized beta and p value, with Benjamini-Hochberg FDR < 0.10 as the significance level.

For the normalized expression data, moduleEigengenes of WGCNA was used to calculate the MEs for the gene modules of interest for the HC and RA after treatment population samples. The MEs for all of the samples were almost completely the same as those for the RA before treatment population (Pearson’s correlation coefficient, 0.99).

An area under the curve (AUC) analysis was performed using pROC v1.15.3 (14). WGCNA hub gene network visualization was performed using igraph v1.2.4.2 (https://igraph.org/), and nodes with a Pearson’s correlation coefficient > 0.6 were connected by an edge.

**Estimation of pre-DC and other cell subfractions by deconvolution**

Using the published data of pre-dendritic cells (pre-DC) subfractions noted by See P *et al.* (GSE80171) (15), our pDC RNA-seq data, which included the treatment resistance modules, were deconvoluted using CIBERSORTx (16). Deconvolution is a machine learning technique for predicting a specific proportion of cells in a bulk sample based on the gene expression profiles specific to the individual cell types, without actually sorting the cells. For the AS DC fraction noted by Villani AC *et al.* (17), as well, using the published data (GSE94820), our RNA-seq data were deconvoluted, and the correspondence of the ratios of AS DC in pDC for which high and low pDC module expression levels were predicted were investigated. The genes that were thought to characterize pre-DC in the published report of See P *et al.* (15) – that is, the DEGs compared to pDC, and the genes that were considered characteristic of pre-DC, were considered pre-DC signature genes (online supplementary table 5). Similarly, the genes that characterized the AS DC population in the report published by Villani AC *et al.* (17) were considered AS DC signature genes (online supplementary table 5).

**Exploration of genes associated with the WGCNA module**

The normalized RNA-seq data from all of the RA before and after treatment patients and the HC were used. Genes correlated with the pre-DC ME with a Benjamini-Hochberg FDR < 0.05 based on an empirical Bayesian method and a linear model using limma v3.38.3 were considered genes associated with pre-DC (18). Genes negatively correlated to pre-DC (log fold change < 0) were subjected to MSigDB hallmark gene set enrichment analysis (12). Genes that were contained in either MSigDB HALLMARK_INTERFERON_ALPHA_RESPONSE or HALLMARK_INTERFERON_GAMMA_RESPONSE are shown in volcano plots as interferon (IFN) response genes (figures 4B and 4C).

**Mediation analysis**

The relationships between CD16n Mono IFN-α response signature expression, pDC_M18, and CDAI50 at 6 months were tested by mediation analysis using R mediation package v4.5.0. The achievement of CDAI50 at 6 months was treated as the dependent variable of the logistic regression model. CD16n Mono IFN-α response signature expression was treated as the independent variable and pDC_M18 was treated as the mediator. P values were calculated via 1000-time bootstrapping.

**Confirmatory cohort 1, assessment of pDC treatment-resistant genes by quantitative PCR (qPCR)**

Adult Japanese RA patients being treated on an outpatient basis by the Allergy and Rheumatology Department of the University of Tokyo Hospital were used as a confirmatory cohort. In a prospective study, peripheral blood samples were collected from RA patients satisfying the ACR/EULAR RA classification criteria 2010 (1) before being started on a new therapy.

This study was approved by the Ethics Committees of the University of Tokyo (G10095). Written informed consent was obtained from each subject in accordance with the Declaration of Helsinki.

PBMCs collected in the same manner as for Datasets 1, 2, and 3, and a MoFlo XDP (Beckman Coulter) was used to sort the pDC by flow cytometry. Sorting was performed with 5,000 cells serving as the upper limit, and the resulting products were stored in a deep freezer at -80$℃$.

Total RNA was extracted with the RNeasy Micro Kit (Qiagen) and was reverse-transcribed to cDNA with random primers (Invitrogen), dNTP mixture (Takara), ribonuclease inhibitor (Promega), and SuperScript Ⅲ (Invitrogen). Quantitative PCR (qPCR) was performed using CFX Connect Real-Time PCR Detection System (Bio-Rad) with QuantiTect SYBR Green PCR Kit (Qiagen). The primer pairs used in this study are shown in online supplementary table 14. Relative expression was calculated based on the abundance of control ACTB.

The expression levels of the top 15 hub genes in pDC_M18 were evaluated. The mean of the Z scores of the expression levels for each gene for each patient was used as the signature.

**Confirmatory Cohort 2, mass cytometric analysis of peripheral blood of patients starting abatacept therapy**

A prospective study, named “PREDICTABA study”, was conducted in adult Japanese RA patients being treated on an outpatient basis by the Allergy and Rheumatology Department of the University of Tokyo Hospital and by 4 other partner research institutions (Department of Allergy and Immunological Diseases at Tokyo Metropolitan Cancer and Infectious Diseases Center Komagome Hospital, Department of Rheumatology at the Tokyo Metropolitan Bokutoh Hospital, Department of Medicine and Rheumatology at the Tokyo Metropolitan Geriatric Medical Center, and the Azuma Rheumatology Clinic) meeting the 2010 ACR/EULAR classification criteria for RA (1) who were going to start receiving treatment with ABT on the basis of a routine medical assessment because they had not responded adequately to one or more csDMARDs, but who had received 3 or fewer biologic drug products (UMIN-CTR study ID: UMIN000041754). Persons with active infections or malignant tumors or a history of hypersensitivity to ABT, as well as pregnant or breastfeeding women or women wanting to conceive, were excluded from the study.

This study was approved by the Ethics Committees of the University of Tokyo (2019017G). Written informed consent was obtained from each subject in accordance with the Declaration of Helsinki.

Peripheral blood samples were collected from the patients, and PBMCs isolated using Lymphoprep (Abbott Diagnostics Technologies AS) were re-suspended in PBS (-) and stained using Cell-ID Cisplatin-198Pt (Fluidigm) for dead cell identification. After the cells were washed twice with Maxpar Cell Staining Buffer (Fluidigm), Human TruStain FcX (Fc Receptor Blocking Solution) (BioLegend) was added to block antibody non-specific binding, and a reaction was allowed to occur for 10 minutes at room temperature. Staining was performed using 36 types of labelled antibodies (online supplementary table 15). An antibody mixture was added and allowed to react for 30 minutes at room temperature, and the resulting product was then washed using Maxpar Cell Staining Buffer (Fluidigm). The cells were then suspended using Maxpar Fix and Perm Buffer (Fluidigm) that contained Cell-ID Intercalator-Ir (Fluidigm), and the cells were then fixed and stained. The resulting product was then washed using Maxpar Cell Staining Buffer (Fluidigm), and then washed twice with Maxpar Water (Fluidigm). The cells were filtered with a 35 μm mesh filter and 10% of EQ Four Element Calibration Beads (Fluidigm) was added. Measurement was performed using Helios (Fluidigm) with capture events 250-300 /sec and raw data were EQ-Bead-normalized using CyTOF Software v7.0.8493.

**Data analysis of immune cell populations by mass cytometry**

Mass cytometric data were loaded into R using flowCORE v2.4.0. Debris and dead cells were manually removed by Cell-ID Intercalator-Ir stainings. From each sample, 100,000 events were saved in an FCS file. A search was then performed for clusters of immune cells associated with treatment resistance using cytometry by time of flight (CyTOF) workflow (19). First, the marker expressions were analyzed using CATALYST v1.16.2 to perform arcsinh transformation with a cofactor of 5. Of the 28 samples analyzed, 1 sample had poor CD4 staining, and was therefore excluded from the analysis. Cell population identification was performed with FlowSOM v2.1.31 (20) and ConsensusClusterPlus v1.50.0 (21). First, a self-organizing map (SOM), in which cells are assigned according to their similarities to 100 grid points of the SOM and there is metaclustering of the SOM grid points with 30 maxK clusters, was built. Three clusters were merged because of the similarity of the expression markers, and 27 immune cell population clusters were therefore ultimately identified. The standardization procedure of the Human Immunology Project was followed (22), and each population was annotated as, for example, “Naïve B,” based on the similarity of the expression markers of each cell population. A differential analysis of responders and non-responders was performed using diffcyt v1.12.0 (23). A generalized linear mixed model (GLMM), where the fixed effect was defined by the treatment response and the random effect was defined by the sample ID, was used to test the differential abundance of each cell population. Finally, 500 cells from each sample were collected, and tSNE was used to perform dimension reduction on the expression data, and the results presented graphically.

**Manual gating evaluation of pre-DC by mass cytometry**

The pre-DC was gated manually using FlowJo (https://www.flowjo.com/solutions/flowjo) as Lin (CD34, CD4, CD14, CD19, CD16)^-^ HLA-DR^+^ CD45RA^+^ CD123^+^ CD11c^nega/mid^ CX3CR1^+^, based on the report published by See P *et al.* (15) (online supplementary figure 8). Specifically, cell populations were identified based on protein expression at the single cell level, and the relationship between the proportion of cells relative to the parent population and resistance to ABT treatment was evaluated.

Additionally, because CD33, one of the markers of pre-DC, was not included in the mass cytometric analysis panel, flow cytometry was used to re-assess the consistency of the pre-DC gating with the pre-DC gating described in the report of See P *et al.* (15). PBMCs were obtained from the blood of HC using the same procedure as that described above, and were analyzed using FACS Aria^TM^ Fusion (BD Biosciences). The fluorescently labelled antibodies shown in online supplementary table 16 were used to stain the PBMCs, and we performed manual gating, defining Lin (CD34, CD3, CD14, CD19, CD16)^–^ HLA-DR^+^ CD45RA^+^ CD123^+^ CD11c^nega/mid^ CX3CR1^+^ as pre-DC (online supplementary figure 8). It was confirmed that 73.8% of the pre-DC as described in the paper of See P *et al.* (15) (Lin(CD34, CD3, CD14, CD19, CD16)^-^ HLA-DR^+^ CD33^+^ CD45RA^+^ CD123^+^), were included in the aforementioned pre-DC gating (online supplementary figure 9).

**Synovial single cell RNA-seq data analysis**

We reanalyzed the single cell RNA-seq data of CD45^+^ immune cells derived from synovial membrane of treatment naïve RA patients (n=16) described previously (24). Briefly, single cell 3’ gene expression libraries were prepared with Chromium Single Cell 3’v2 Library kit (10× Genomics), and sequencing was performed with Illumina Xten PE-150 platform (ANOROAD and Novogene). The RNA-seq data were aligned to human GRCh38 genome. FindNeighbors and FindClusters function of Seurat v3.1.1 (25) identified 20 major synovial immune clusters using K-nearest neighbor graph based clustering and Louvain algorithm, as reported previously (24). We selected 3804 DC cells from four DC clusters (cDC, S100B^hi^ DC, LAMP3 DC, and pDC) for further analysis. We reclustered the synovial DC data with Seurat’s FindNeighbors and FindClusters in different number of input principal components (PCs) and different resolution parameters, to optimize and characterize DC subpopulation enriched with pre-DC related “pDC_M18 gene module” (online supplementary table 4). As a result, clustering analysis with 10 PCs and resolution parameter 0.3 identified 10 DC clusters, that were visualized using UMAP. Differentially overexpressed genes of DC clusters were identified with FindAllMarkers function of Seurat with default parameters. Each synovial DC clusters were renamed, such as LAMP3^+^ DC, based on their marker gene expression. Based on the distribution of disease duration, the patients with disease duration < 3 years (n=9, “Early” patients) and those patients with disease duration > 3 years (n=7, “Late” patients) were compared for DC cluster frequencies.

To biologically characterize differentially overexpressed genes in DC clusters, enrichment of overexpressed genes of cDC1 and cDC2 populations was analyzed. From original differentially expressed gene table by See *et al.* (15) (table S5), we used genes differentially overexpressed only in cDC1 or cDC2. For the enrichment analysis of anti-inflammatory cDC2a and pro-inflammatory cDC2b gene analysis, we used core gene list of mouse cDC2s by Brown *et al.* (26) (table S3). Gene names were converted to human ortholog by biomaRt v2.52.0.

**Statistical analysis**

R v3.5.1 and R v4.1.0 (R Foundation for Statistical Computing) were used for statistical analysis. Categorical data were assessed using Fisher’s exact probability test. For intergroup testing of quantitative variables, the Shapiro-Wilk test was performed to check for the presence or absence of normality, and the Mann-Whitney U test was performed if the distribution was not a normal distribution. If the distribution was a normal distribution, then Student’s t-test was performed if there was homogeneity of variance on an F-test, and Welch’s t-test was performed if there was not homogeneity of variance.

Spearman’s rank correlation coefficient was used for the correlation analysis if the distribution was not a normal distribution, and Pearson’s correlation coefficient was used if the distribution was a normal distribution.

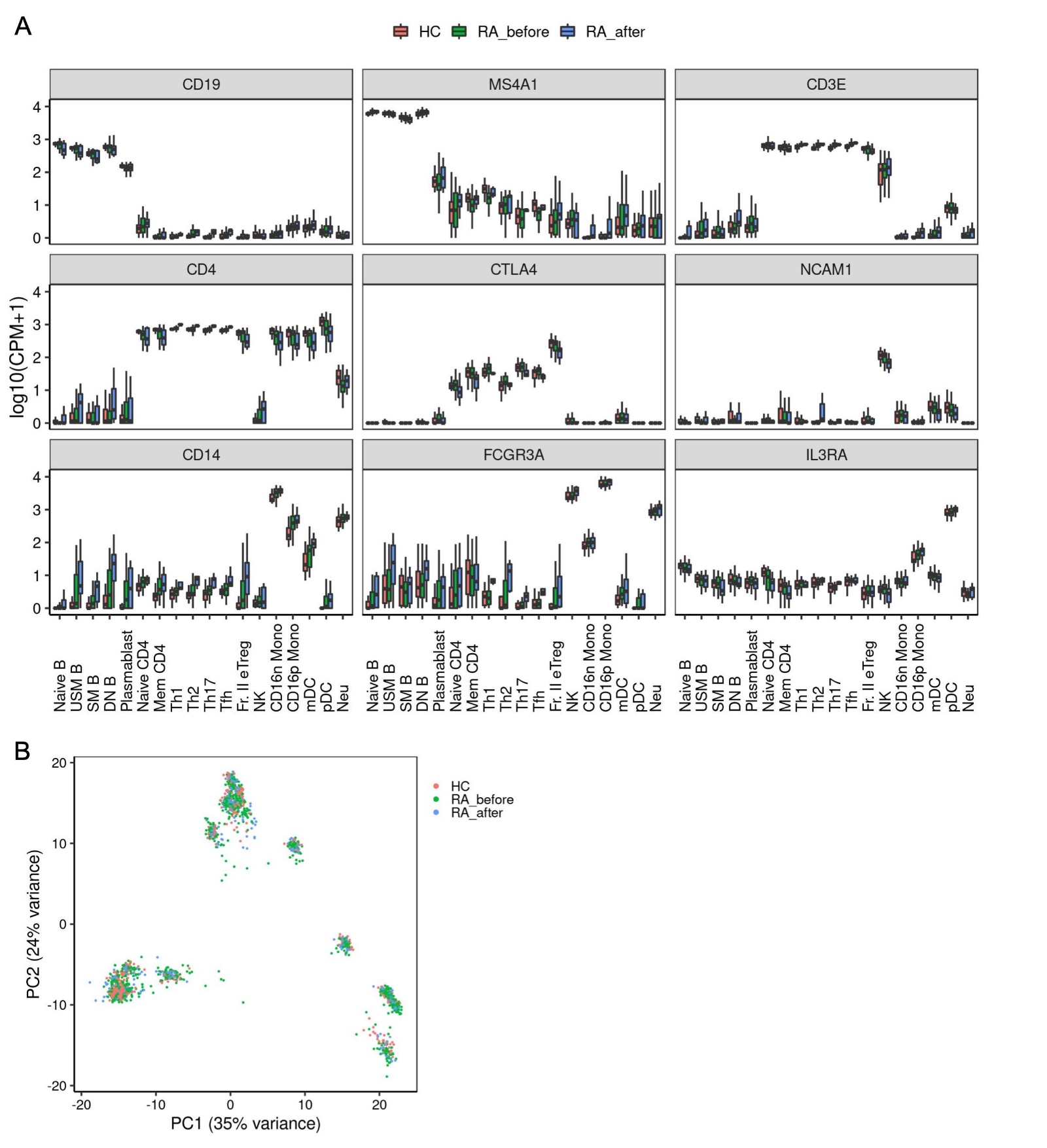

**Online supplementary figure 1. Validity of immune cell RNA-seq data.**

(A) Expression levels of typical genes characteristically highly expressed by each type of cell in the RNA-seq data. (B) Comparison of the HC, RA before treatment, and RA after treatment samples in RNA-seq PCA.

HC; healthy control, RA; rheumatoid arthritis, RNA-seq; RNA-sequencing, PCA; principal component analysis.

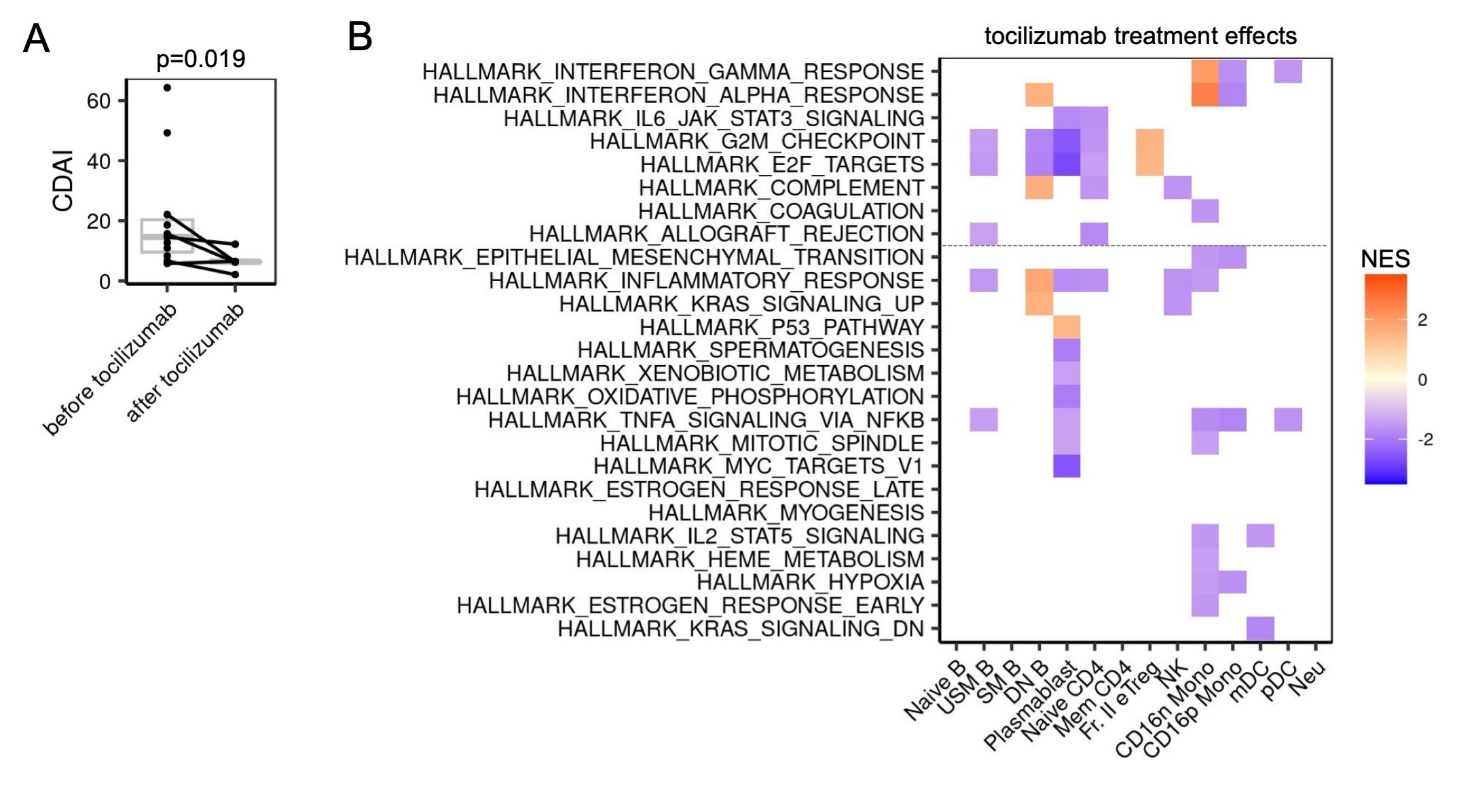

**Online supplementary figure 2. Treatment effects of tocilizumab on immune cell gene expression.**

(A) Clinical treatment effects of tocilizumab. (B) The GSEA results for RA before and after treated with tocilizumab. The 8 gene sets from (figure 2A) with increased expression in the RA population and the gene sets with a change in the |NES| > 1 in at least 1 subset are shown. Pathways with FDR < 0.05 are colored.

NES: normalized enrichment score, GSEA: gene set enrichment analysis. Definitions of the subsets are provided in online supplementary table 2.

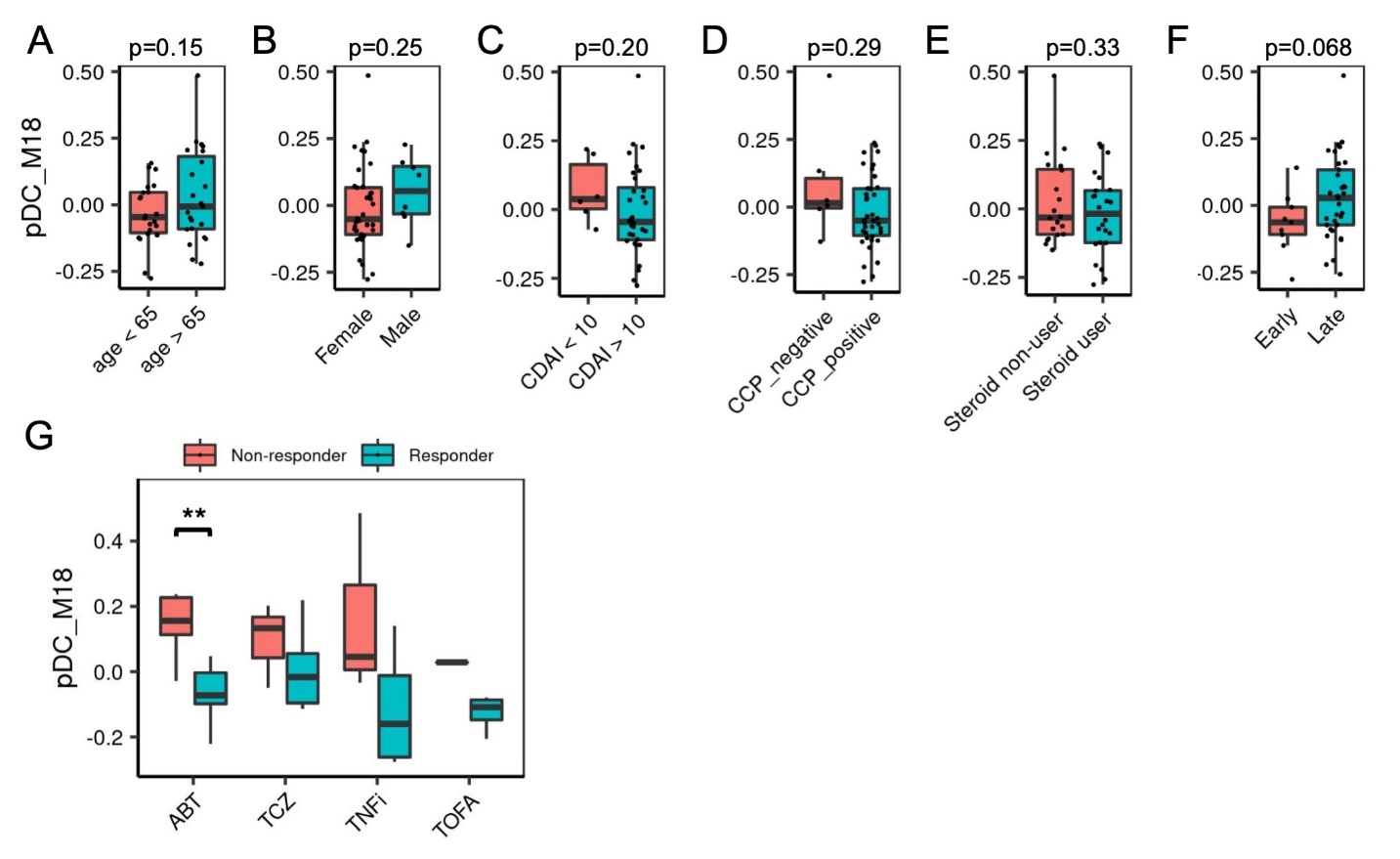

**Online supplementary figure 3. Correlation of clinical measures to pDC_M18.**

The relationships to differences in expression levels in pDC_M18 were evaluated with the analyzed patients divided into 2 groups based on clinical characteristics. (A) age at blood sample collection, (B) sex, (C) CDAI at blood sample collection, (D) ACPA positive/negative, (E) steroid user or non-user, (F) Duration of illness (duration < 1 year = Early), (G) relationship of therapeutic medication and pDC_M18. **p < 0.01.

CCP; cyclic citrullinated peptides, ABT; abatacept, TCZ; tocilizumab, TNFi; tumor necrosis factor inhibitor, TOFA; tofacitinib.

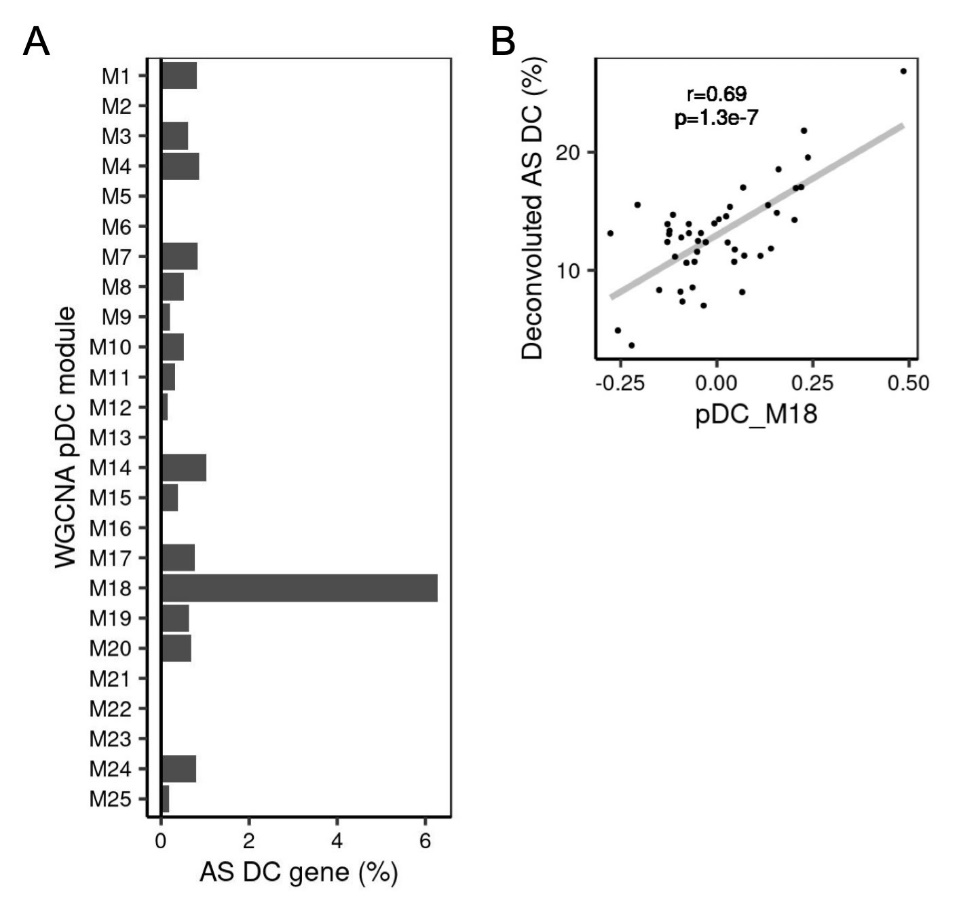

**Online supplementary figure 4. The treatment resistance pDC_M18 module reflects the proportion of AS DC in pDC.**

(A) The percent match with AS DC signature genes was assessed for 25 pDC modules. (B) The correlation of the pDC_M18 ME with the ratio of pre-DC deconvoluted based on the method described in the report of Villani AC *et al.* (17).

WGCNA; weighted gene co-expression network analysis.

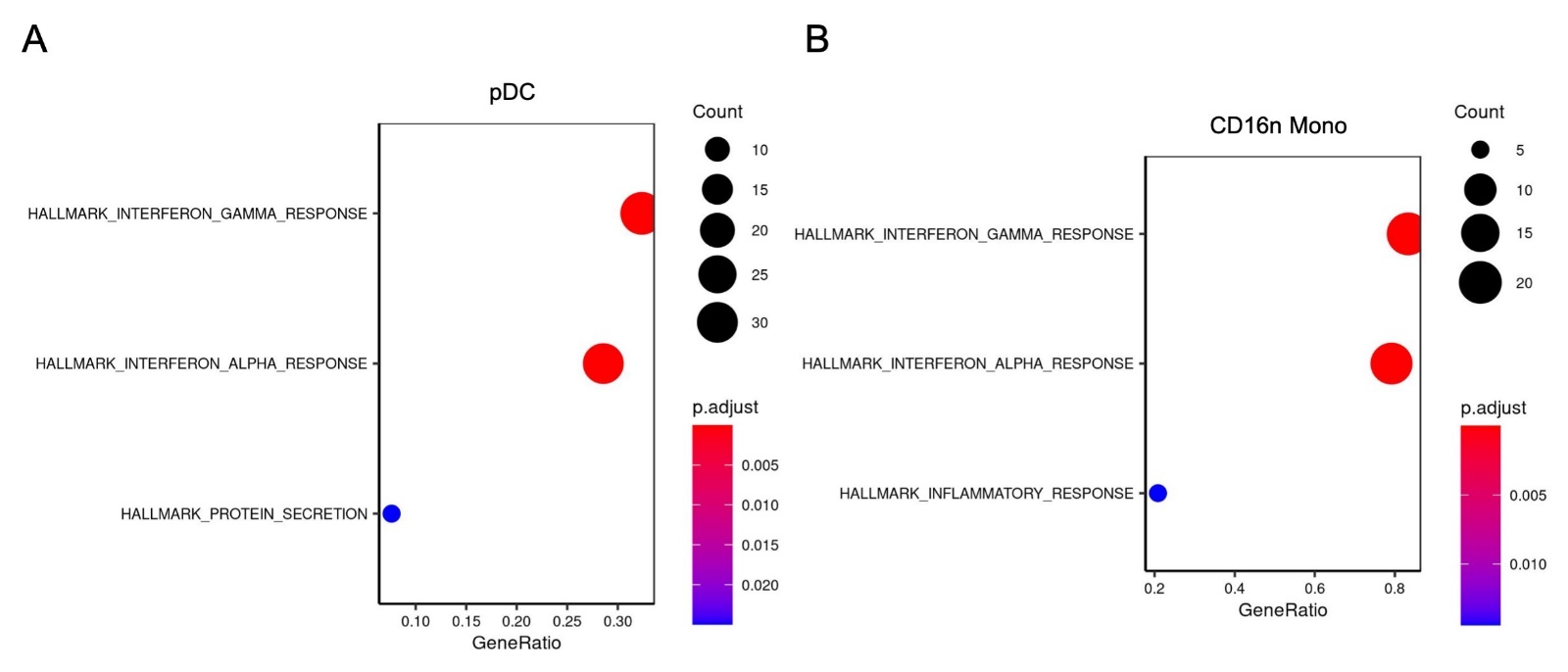

**Online supplementary figure 5. Enrichment analysis of genes negatively correlated to pDC_M18.**

Agreement with the MSigDB Hallmark pathway of genes negatively correlated to pDC_M18 in pDC (A) and CD16n Mono (B).

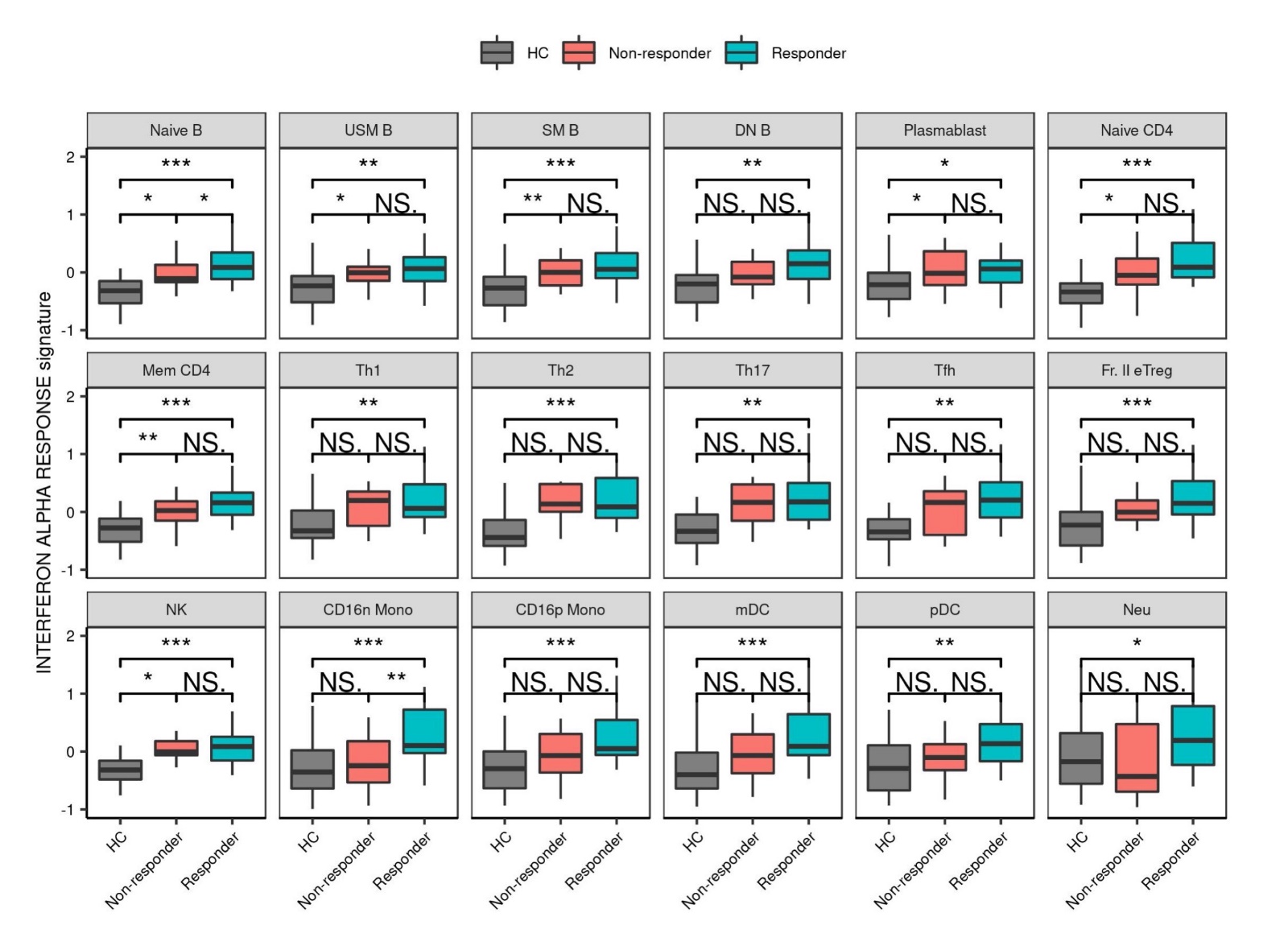

**Online supplementary figure 6. IFNα response signature and RA prognosis.**

The IFNα response signatures of the HC population and the RA before treatment population, stratified by treatment prognosis, were compared. *p < 0.05, **p < 0.01, ***p < 0.001.

HC; healthy control, RA; rheumatoid arthritis, NS; not significant, IFN; interferon.

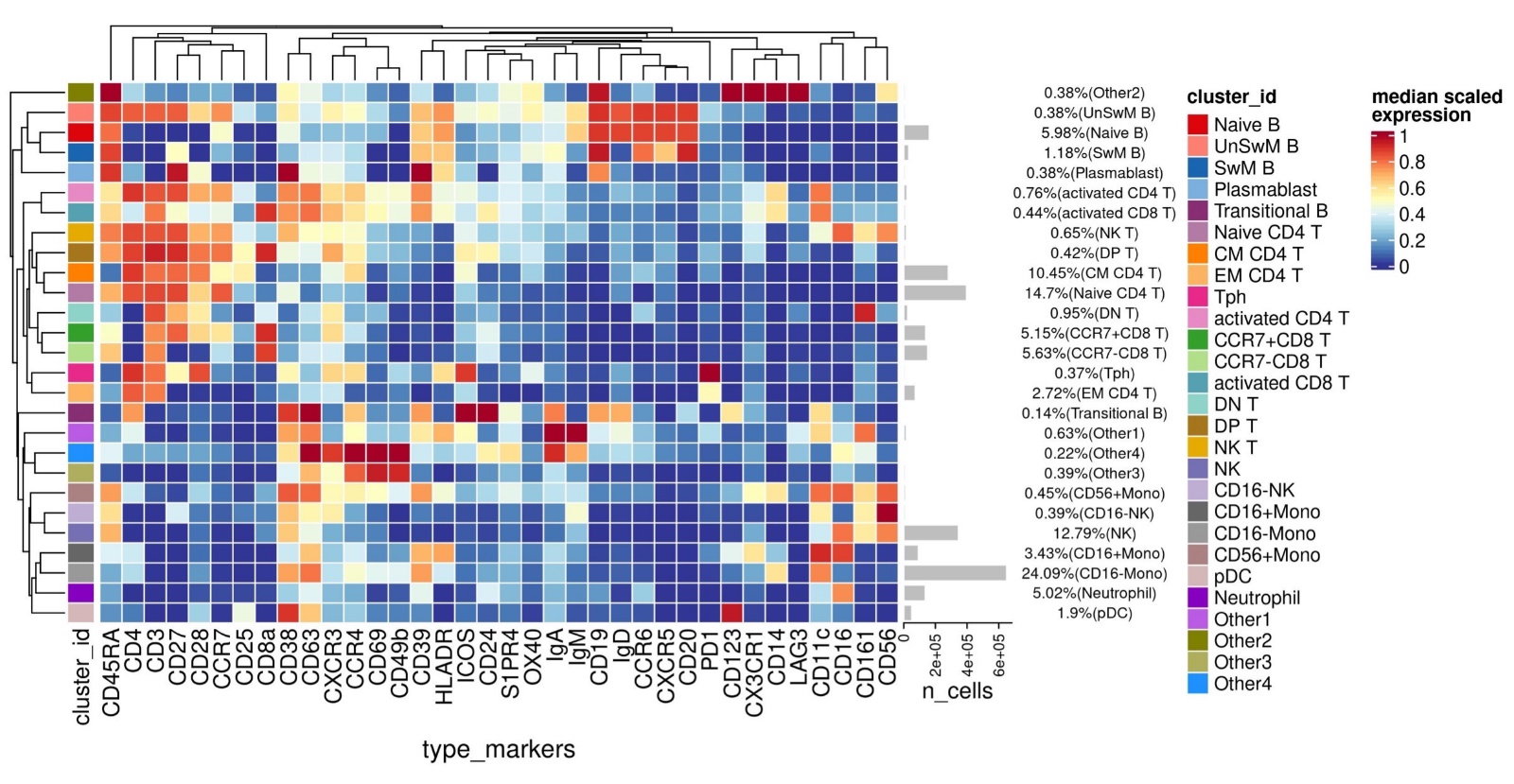

**Online supplementary figure 7. Identification of immune cell populations by mass cytometry.**

The expression of the 36 cell surface markers in 27 cell population clusters were scaled to create a cluster heat map. The numbers and proportions of cells in each cell population cluster are shown on the right. The names of the 27 cell population clusters were determined based on their expression of the cell surface markers.

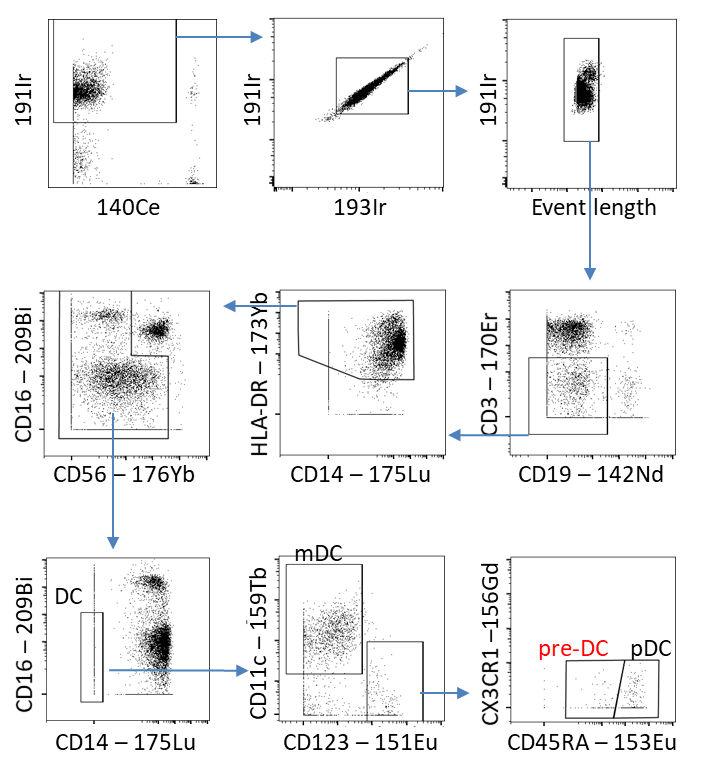

**Online supplementary figure 8. pre-DC gating in the confirmatory cohort analyzed by mass cytometry.**

The mass cytometric data obtained from the peripheral blood samples collected before treatment from 28 patients in the confirmatory cohort who had been treated with abatacept (PREDICTABA study) was used to gate the pre-DC as Lin(CD34, CD3, CD14, CD19, CD16)^-^ HLA-DR^+^ CD45RA^+^ CD123^+^ CD11c^nega/mid^ CX3CR1^+^.

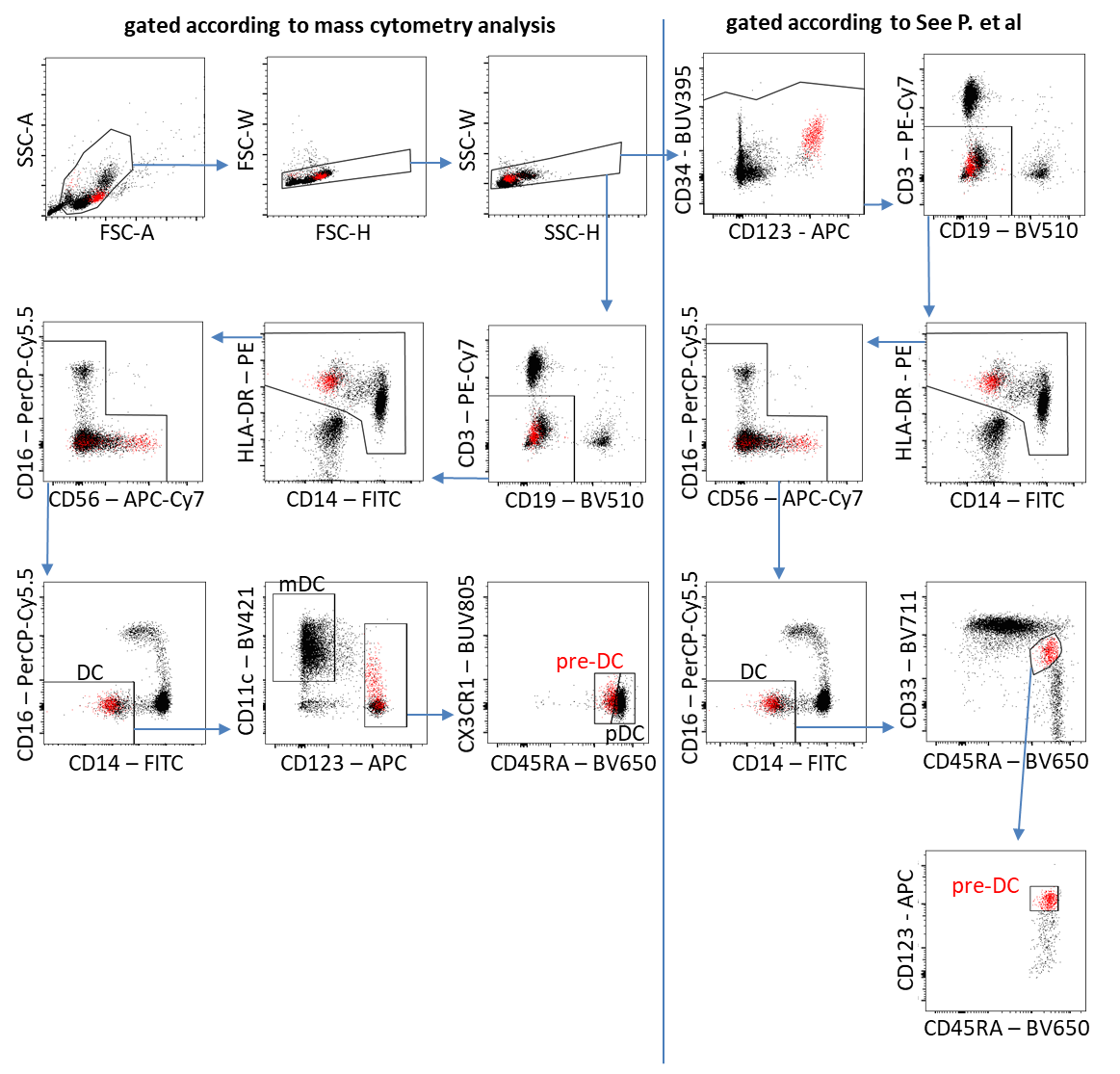

**Online supplementary figure 9. Assessment of the probability of pre-DC gating in mass cytometry analysis.**

Flow cytometry was used to assess whether or not the results of gating pre-DC in accordance with the mass cytometric analysis procedure used for our confirmatory cohort (Lin(CD34, CD3, CD14, CD19, CD16)^-^ HLA-DR^+^ CD45RA^+^ CD123^+^ CD11c^nega/mid^ CX3CR1^+^) were consistent with the results obtained by gating the pre-DC based on the procedure described in the report of See P *et al.* (15) (Lin(CD34, CD3, CD14, CD19, CD16)^-^ HLA-DR^+^ CD33^+^ CD45RA^+^ CD123^+^).

**Online supplementary table 1. Clinical background information for the RA and HC populations.**

|  |  | RA (n=55) | HC (n=39) | p-value |
| --- | --- | --- | --- | --- |
| male/female, n (%) | | 10/45 (18.2/81.8) | 6/33 (15.4/84.6) | 0.79 |
| age, years, median (IQR) | | 62.7 (55.0-71.5) | 61.0 (51.5-70.0) | 0.51 |
| disease duration, years, median (IQR) | | 10.0 (1.3-16) |  |  |
| interstitial pneumonia +/-, n (%) | | 6/49 (11.9/89.1) |  |  |
| before new treatment | ACPA +/-, n (%) | 47/7 (85.5/12.7) |  |  |
|  | RF+/-, n (%) | 47/8 (85.5/14.5) |  |  |
|  | CRP, mg/dl, median (IQR) | 2.2 (0.4-3.6) |  |  |
|  | CDAI, median (IQR) | 23.3 (12.1-36.3) |  |  |
| 6 months after the new treatment | CDAI, median (IQR) | 11.8 (3.8-16.3) |  |  |
|  | CDAI50 archivement/non-archivement, n(%) | 27/18 (49.1/32.7) |  |  |
|  | EULAR response criteria | Good 15, Moderate18, Poor 13, Unknown 9 |  |  |
| new treatment | | ABT 22, TCZ 12, TOFA 6, MTX 6, GLM 4, ADA 1, ETN 1, IFX 1, IGU 1, SASP 1 |  |  |
| second RNA-seq after treatment | | ABT 15, TCZ 5 |  |  |
| RA, rheumatoid arthritis; HC, healthy control; IQR, interquartile range; ACPA, anticyclic citrullinated peptide antibodies; RF, rheumatoid factor; CRP, C-reactive protein; EULAR, European League Against Rheumatism; ABT, abatacept; TCZ, tocilizumab; TOFA, tofacitinib; MTX, methotrexate; GLM, golimumab; ADA, adalimumab; ETN, etanercept; IFX, infliximab; IGU, iguratimod; SASP, salazosulfapyridine. | | | | |

**Online supplementary table 2. Peripheral blood immune cell subsets analyzed, and definitions thereof.**

| Subset name | Abbreviation | Definition (Dataset 1-3) | Definition (Dataset 4) |
| --- | --- | --- | --- |
| **CD4 T cells** | CD4 |  |  |
| Naïve CD4 T cells | Naïve CD4 | CD3^+^CD4^+^/CD25^-^/CD45RA^+^ | CD3^+^/CD4^+^CD8^-^/CCR7+CD45RA+ |
| Memory CD4 T cells | Mem CD4 | CD3^+^CD4^+^/CD25^-^/CD45RA^-^ | CD3^+^/CD4^+^CD8^-^/non-naive CD4^+^/CD25^-^ |
| T helper 1 cells | Th1 | CD3^+^CD4^+^/CD25^-^/CD45RA^-^CXCR5^-^/CCR6^-^CXCR3^+^ | CD3^+^/CD4^+^CD8^-^/non-naive CD4^+^/CD25^-^/CXCR5^-^CCR6^-^/CXCR3^+^CCR4^-^ |
| T helper 2 cells | Th2 | CD3^+^CD4^+^/CD25^-^/CD45RA^-^CXCR5^-^/CCR6^-^CXCR3^-^ | CD3^+^/CD4^+^CD8^-^/non-naive CD4^+^/CD25^-^/CXCR5^-^CCR6^-^/CXCR3^-^CCR4^+^ |
| T helper 17 cells | Th17 | CD3^+^CD4^+^/CD25^-^/CD45RA^-^CXCR5^-^/CCR6^+^CXCR3^-^ | CD3^+^/CD4^+^CD8^-^/non-naive CD4^+^/CD25^-^/CXCR5^-^CCR6^+^/CXCR3^-^ |
| T follicular helper cells | Tfh | CD3^+^CD4^+^/CD25^-^/CD45RA^-^CXCR5^+^ | CD3^+^/CD4^+^CD8^-^/non-naive CD4^+^/CD25^-^/CXCR5^+^ |
| Fraction II effector regulatory T cells | Fr. II eTreg | CD3^+^CD4^+^/CD25^++^LAG3^-^/CD45RA^-^ | CD3^+^/CD4^+^CD8^-^/CD25^++^CD45RA^-^ |
| **B cells** | B |  |  |
| Naïve B cells | Naïve B | CD3^-^CD19^+^/IgD^+^CD27^-^ | CD3^-^CD19^+^/IgD^+^CD27^-^ |
| Unswitched  memory B cells | USM B | CD3^-^CD19^+^/IgD^+^CD27^+^ | CD3^-^CD19^+^/IgD^+^CD27^+^ |
| Switched memory  B cells | SM B | CD3^-^CD19^+^/IgD^-^CD27^+^ | CD3^-^CD19^+^/IgD^-^CD27^+^/CD38^-^ |
| Double Negative  B cells | DN B | CD3^-^CD19^+^/IgD^-^CD27^-^ | CD3^-^CD19^+^/IgD^-^CD27^-^ |
| Plasmablasts | Plasmablast | CD3^-^CD19^+^/IgD^-^/CD27^++^CD38^+^ | CD3^-^CD19^+^/IgD^-^CD27^++^/CD38^+^ |
| **Natural Killer cells** | NK | CD3^-^CD19-/CD14-/CD56+ | CD3^-^CD19^-^/CD14^-^/CD56^+^ |
| **Monocytes** | Mono |  |  |
| CD16 positive  monocytes | CD16p Mono | CD3^-^CD19^-^/HLADR^+^/CD56^-^/CD14^+^CD16^+^ | CD3^-^CD19^-^/HLA-DR^+^/CD56^-^/CD14^+^CD16^+^ |
| CD16 negative  monocytes | CD16n Mono | CD3^-^CD19^-^/HLADR^+^/CD56^-^/CD14^+^CD16^-^ | CD3^-^CD19^-^/HLA-DR^+^/CD56^-^/CD14^+^CD16^-^ |
| **Dendric cells** | DC |  |  |
| Myeloid dendritic  cells | mDC | CD3^-^CD19^-^/CD56^-^CD16^-^/HLADR^+^CD14^-^/CD11c^+^CD123^-^ | CD3^-^CD19^-^/HLA-DR^+^/CD56^-^/CD14^-^CD16^-^/CD11c^+^CD123^-^ |
| Plasmacytoid  dendritic cells | pDC | CD3^-^CD19^-^/CD56^-^CD16^-^/HLADR^+^CD14^-^/CD11c^-^CD123^+^ | CD3^-^CD19^-^/HLA-DR^+^/CD56^-^/CD14^-^CD16^-^/CD11c^-^CD123^+^ |
| **Neutrophils** | Neu | Immune-magnetically sorting with the "EasySep direct human neutrophil isolation kit" | Immune-magnetically sorting with the "MACSxpress Neutrophil isolation Kit, human" |

**Online supplementary table 3. Clinical information on the responder and non-responder populations.**

|  |  | Responder (n=27) | Non-responder (n=18) | p-value |
| --- | --- | --- | --- | --- |
| male/female, n (%) | | 3/24 (11.1/88.9) | 4/14 (22.2/77.8) | 0.41 |
| age, years, median (IQR) | | 61.1 (55-69.5) | 63.6 (56.5-72.5) | 0.66 |
| Dataset 1/2/3/4, n (%) | | 10/3/2/12 (37.0/11.1/7.41/44.4) | 11/2/1/4 (61.1/11.1/5.6/22.2) |  |
| disease duration, years, median (IQR) | | 12.5 (6.0-19.0) | 8.5 (2.25-8) | 0.16 |
| interstitial pneumonia +/-, n (%) | | 2/25 (7.4/92.6) | 4/14 (22.2/77.8) | 0.2 |
| before new treatment | ACPA +/-, n (%) | 24/2 (88.9/7.4) | 15/3 (83.3/16.7) | 0.39 |
|  | RF +/-, n (%) | 24/3 (88.9/11.1) | 15/2 (83.3/11.1) | >0.99 |
|  | CRP, mg/dl, median (IQR) | 2.3 (0.6-3.6) | 2.5 (0.4-5.2) | 0.89 |
|  | CDAI, median (IQR) | 25.0 (13,4-38.5) | 21.1 (12.5-33.4) | 0.46 |
|  | PSL, mg/day, median (IQR) | 2.4 (0-4.0) | 3.1 (0-5.8) | 0.7 |
|  | MTX, +/-, n (%) | 17/10 (63.0/27.0) | 6/12 (33.3/66.7) | 0.071 |
| 6 months after the new treatment | CDAI, median (IQR) | 7.4 (2.9-10.5) | 18.5 (12.4-28.0) | 0.00066 * |
|  | EULAR response criteria | Good 13, Moderate 13, Poor 1 | Good 2, Moderate 4, Poor 12 |  |
| new treatment | details | ABT / TCZ 8, TOFA 5, GLM 2, IFX / ADA / MTX / SASP 1 | ABT 10, TCZ 3, GLM 2, TOFA / ETN / IGU 1 |  |
|  | bio switch +/-, n (%) | 5/22 (18.5/81.5) | 4/14 (22.2/77.8) | >0.99 |
| IQR, interquartile range; ACPA, anticyclic citrullinated peptide antibodies; RF, rheumatoid factor; CRP, C-reactive protein; PSL, prednisolone; MTX, methotrexate; EULAR, European League Against Rheumatism; ABT, abatacept; TCZ, tocilizumab; TOFA, tofacitinib; GLM, golimumab; IFX, infliximab; ADA, adalimumab; SASP, salazosulfapyridine; ETN, etanercept; IGU, iguratimod. | | | | |
| *p < 0.05 | | | | |

**Online supplementary table 4. Hub genes in pDC_M18, the module prepared by WGCNA.**

pDC_M18

COTL1 IFI30 FGL2 ABI3 CX3CR1 LILRA2 GFRA2 KLF4 CYP2S1 CD22 TIMP1 S100A10 SLC24A4 CEBPD NFE2 APOL3 CFP SPI1 MS4A7 CD33 CEACAM4 CLEC10A IL1RN RTN1 LPCAT2 ATP1A2 MBOAT2 LAT2 NAIP RIN3 ADORA2B CD300C LGALS3 CEBPA FBLN2 IGFBP7 HAMP FGR TCEA3 KCNK6 TIAM1 ASGR2 CES1 ARRB1 AXL GSE1 HSPA12B SIGLEC6 IL1B TM6SF1 TNFAIP2 ZNF366 PADI2 GSAP C20orf27 GPR146 CXCR2 ADAM33 GAS7 IL7 CD244 FAM129A DGAT2 TNFRSF25 CFD BASP1 TLR4 GPAT3 CD151 ITGB2.AS1 AIM1 CARD9 SIGLEC1 DENND3 LYZ DOK2 FCGRT UBASH3B RGCC TSPAN32 SLC46A2 KIT CSTA CPXM1 HAVCR2 GLIPR2 SEMA4B ASL SULT1B1 EMP1 LILRA1 PLXDC2 ACPP EHD3 DHRS3 ADAP1 SLCO3A1 TSPAN15 LIMA1 CPNE2 GPR35 RALB ULK2 LDLRAP1 CLIC2 ANXA1 MAGEF1 SPOCK2 ASAP1 ASCL2 PIK3R6 MARVELD1 CD163 PHLDA3 STMN2 ALOX5 LRP1 PTGS1 PLTP SNX30 NUDT16P1 ZNF385A PTK6 FAM46C SIRPB2 ITGAX COL9A2 AHR GAS1 PLBD1 ANPEP RGS16 NOL4L SCIMP B4GALT2 HIP1 METRNL VIPR1 RXRA KLF8 LPAR5 CBL SLC2A3 ZBTB7B BCL6 IRAK3 TLE4 MTMR11 FAM198B SPECC1 SEMA4C RIBC2 UNC5CL CAMP HMGB3 NCAPH NT5DC2 RARG SNX33 RASGRP4 APAF1 CST7 RAB32 APBB1 CTSH RHOU EFNB1 CSRP1 CLEC12A ZAK SH3RF1 ZNF324 PPARGC1B SLC9A9 ANXA2 ADGRG1 G6PC3 CD93 PSTPIP2 CD63 PROCR CRK C11orf21 CASP1 CAMKK2 CREB5 ATG7 ZNF69 AMPD3 C19orf38 XYLT1 TUBA4A PSEN2 HAL C7orf50 RARA.AS1 MYADM ADAM28 MSLN SDPR TBL1X SPRY2 LCP2 PRF1 ARID5B CD200R1 GNGT2

WGCNA, weighted gene co-expression network analysis.

**Online supplementary table 5. List of previously reported pre-DC signature genes (15) and AS DC signature genes (17).**

pre-DC signature genes (See P *et al*. Science 2017)

BTLA CD48 CD2 CD22 CD244 CD33 CD44 CD45RA CD5 CD63 CD72 CD86 CD93 CD141 CD303 CD304 CLEC10A CLEC12A CLEC4A CSF3R CX3CR1 CXCL16 HLA-DRA ICAM3 ICAM4 IFITM2 IFITM3 IL13RA1 IL1R2 IRF4 IRF8 ITGA5 ITGAX KLF4 KLF8 LILRB1 LILRB3 LTBR NLRP3 PACSIN2 RAB31 RAB32 RAB7A RUNX3 SIGLEC10 SIGLEC6 THBD TNFSF12 ZBTB46

AS DC signature genes (Villani AC *et al*. Science 2017)

ACPP ADAM33 AK125727 ALDH2 APEX1 ARHGAP18 ATF5 ATP2B4 AXL BAIAP2 BHLHE40 BIN1 C5ORF25 CCND3 CD22 CD300LB CD300LG CD5 CD72 CDH1 CDKN1A CEP95 COQ7 CTSW CX3CR1 CXCR2 DAB2 DPYSL2 ENTPD7 FAM105A FAM129A GGTA1P GNAQ GPR146 HIP1 IRF4 KLF12 KLF4 LAX1 LGMN LTK MECR MED12L MGLL MRPS6 MYH11 MYO1E NDRG1 PIM2 PLA2G16 PLAC8 PPP1R14A PTGDS RAD1 RBL1 RNASEL RNF141 RUNX2 S100A10 SCN9A SEPT6' SIGLEC1 SIGLEC6 SLC20A1 SLC35C2 SLC41A2 SLC4A3 SNRNP25 SOX4 SP4 STAG3L4 STX18 SUCLA2 SUSD1 TBC1D9 THBD TNFSF12 TNNI2 TSEN54 TXN UPK3A USF2 VASH1 ZEB1 ZNF789

**Online supplementary table 6. Clinical information for the responder and non-responder populations in the confirmatory cohort for which qPCR was performed.**

|  |  | Responder (n=9) | Non-responder (n=10) | p-value |
| --- | --- | --- | --- | --- |
| male/female, n (%) | | 0/9 (0/100.0) | 1/9 (10.0/90.0) | >0.99 |
| age, years, median (IQR) | | 53.7 (38.0-64.0) | 68.9 (60.3-76.3) | 0.048 * |
| disease duration, years, median (IQR) | | 7.5 (3.0-14.3) | 13.0 (6.9-16.9) | 0.13 |
| interstitial pneumonia +/-, n (%) | | 1/8 (11.1/88.9) | 4/6 (40.0/60.0) | 0.3 |
| before new treatment | ACPA +/-, n (%) | 7/2 (77.8/22.2) | 8/2 (80.0/20.0) | >0.99 |
|  | RF +/-, n (%) | 8/1 (88.9/11.1) | 7/3 (70.0/30.0) | 0.58 |
|  | CRP, mg/dl, median (IQR) | 1.2 (0.1-0.7) | 1.1 (0.1-1.5) | 0.54 |
|  | CDAI, median (IQR) | 23.3 (11,5-33.0) | 19.3 (10.5-24.8) | 0.61 |
|  | PSL, mg/day, median (IQR) | 3.2 (0-6.0) | 5.5 (4.3-6.9) | 0.18 |
|  | MTX, +/-, n (%) | 7/2 (77.8/22.2) | 5/5 (50.0/50.0) | 0.35 |
| 6 months after the new treatment | CDAI, median (IQR) | 4.6(1.0-5.5) | 18.5 (11.6-27.0) | 0.0042 * |
|  | EULAR response criteria | Good 5, Moderate 2 | Good 1, Moderate 2, Poor 6 |  |
| new treatment | details | ABT / GLM 3, ETN 2, TCZ 1 | ABT 6, TCZ / GLM /ETN / SAR 1, for each |  |
|  | bio switch +/-, n (%) | 1/8 (11.1/88.9) | 4/6 (40.0/60.0) | 0.3 |
| IQR, interquartile range; ACPA, anticyclic citrullinated peptide antibodies; RF, rheumatoid factor; CRP, C-reactive protein; PSL, prednisolone; MTX, methotrexate; EULAR, European League Against Rheumatism; ABT, abatacept; GLM, golimumab; ETN, etanercept; TCZ, tocilizumab; SAR, sarilumab. | | | | |
| *p < 0.05 | | | | |

**Online supplementary table 7. Clinical information for the responder and non-responder populations in the mass cytometric analysis of peripheral blood from abatacept patients.**

|  |  | Responder (n=21) | Non-responder (n=7) | p-value |
| --- | --- | --- | --- | --- |
| male/female, n (%) | | 9/12 (42.9/57.1) | 3/4 (42.9/57.1) | >0.99 |
| age, years, median (IQR) | | 72.3 (72.0-80.0) | 60.7 (53.0-72.0) | 0.017 * |
| disease duration, years, median (IQR) | | 7.2 (2.0-8.0) | 10.9 (0.0-20.5) | 0.98 |
| interstitial pneumonia +/-, n (%) | | 7/14 (33.3/66.7) | 2/5 (28.6/71.4) | >0.99 |
| before new treatment | ACPA +/-, n (%) | 20/1 (95.2/4.8) | 7/0 (100.0/0) | >0.99 |
|  | RF +/-, n (%) | 19/2 (90.5/9.5) | 7/0 (100.0/0) | >0.99 |
|  | CRP, mg/dl, median (IQR) | 1.7 (0.3-1.9) | 1.0 (0.1-1.7) | 0.3 |
|  | CDAI, median (IQR) | 17.4 (11.0-21.0) | 21.8 (13.6-30.3) | 0.43 |
|  | PSL, mg/day, median (IQR) | 2.5 (0-3.0) | 3.6 (3.0-4.0) | 0.057 |
|  | MTX, +/-, n (%) | 11/10 (52.4/47.6) | 4/4 (50.0/50.0) | >0.99 |
| 6 months after the new treatment | CDAI, median (IQR) | 2.7 (0.2-3.0) | 13.7 (8.4-17.8) | 0.0055 * |
| Inducted ABT as the 3rd/2nd/1st biologics. n (%) | | 1/4/16 (4.8/19.0/76.2) | 0/3/4 (0/42.9/57.1) | 0.14 |
| IQR, interquartile range; ACPA, anticyclic citrullinated peptide antibodies; RF, rheumatoid factor; CRP, C-reactive protein; PSL, prednisolone; MTX, methotrexate; ABT, abatacept. | | | | |
| *p < 0.05 | | | | |

**Online supplementary table 8. Proportions of cells for each subset in the responder and non-responder populations in the mass cytometric analysis of peripheral blood from abatacept patients.**

|  | subset ratio per DC (%) | | |  |
| --- | --- | --- | --- | --- |
| Subset name | Abbreviation | Responder (n=21) | Non-responder (n=7) | p-value |
| **Dendric cells** | DC |  |  |  |
| Myeloid dendritic cells | mDC | 55.4 (49.9-60.1) | 47.2 (40.5-57.5) | 0.27 |
| Plasmacytoid dendritic cells | pDC | 21.4 (16,2-23.3) | 24.0 (20.6-30.8) | 0.25 |
| Dendritic cells precursors | pre-DC | 1.6 (1.2-2.1) | 3.7 (3.0-4.3) | 0.000004* |
| *p < 0.05 | | | | |

**Online supplementary table 9. Features and sample sizes of each dataset.**

|  | Dataset 1 | Dataset 2 | Dataset 3 | Dataset 4 | total |
| --- | --- | --- | --- | --- | --- |
| cell sorter | MoFlo XDP | MoFlo XDP | MoFlo XDP | FACS Aria^TM^ Fusion |  |
| neutrophil isolation kit | EasySep direct human neutrophil isolation kits | MACSxpress Neutrophil isolation Kits | MACSxpress Neutrophil isolation Kits | MACSxpress Neutrophil isolation Kits |  |
| RNA purification kit | NucleoSpin RNA or RNeasy Micro Kit | RNeasy Micro Kit | RNeasy Micro Kit | MagMAXTM 96 Total RNA Isolation Kit |  |
| library preparation | SMART-seqⅤ4 Ultra Low Input RNA kit for Sequencing | | | |  |
| Next Generation Sequencers | HiSeq2500 | NovaSeq6000 | HiSeq2500 | HiSeq2500 |  |
| RA before treatment | 27 | 5 | 5 | 18 | 55 |
| RA after treatment | 15 | 5 | 0 | 0 | 20 |
| HC | 15 | 0 | 0 | 24 | 39 |
| reference | new data | new data | Ota *et al.* Cell 2021 | Ota *et al.* Cell 2021 |  |
| RA, rheumatoid arthritis; HC, healthy control. | | | | | |

**Online supplementary table 10. Definitions of the peripheral blood immune cell subsets in Datasets 1-3.**

| Dataset 1-3 |  |  |
| --- | --- | --- |
| Subset name | Abbreviation | Definition |
| **CD4 T cells** | CD4 |  |
| Naïve CD4 T cells | Naïve CD4 | CD3^+^CD4^+^/CD25^-^/CD45RA^+^ |
| Memory CD4 T cells | Mem CD4 | CD3^+^CD4^+^/CD25^-^/CD45RA^-^ |
| T helper 1 cells | Th1 | CD3^+^CD4^+^/CD25^-^/CD45RA^-^CXCR5^-^/CCR6^-^CXCR3^+^ |
| T helper 2 cells | Th2 | CD3^+^CD4^+^/CD25^-^/CD45RA^-^CXCR5^-^/CCR6^-^CXCR3^-^ |
| T helper 17 cells | Th17 | CD3^+^CD4^+^/CD25^-^/CD45RA^-^CXCR5^-^/CCR6^+^CXCR3^-^ |
| T follicular helper cells | Tfh | CD3^+^CD4^+^/CD25^-^/CD45RA^-^CXCR5^+^ |
| Fraction II effector  regulatory T cells | Fr. II eTreg | CD3^+^CD4^+^/CD25^++^LAG3^-^/CD45RA^-^ |
| **CD8 T cells** | CD8 |  |
| Naïve CD8 T cells | Naïve CD8 | CD3^+^CD19^-^/CD4^-^CD8^+^/CD45RA^+^ |
| Memory CD8 T cells | Mem CD8 | CD3^+^CD19^-^/CD4^-^CD8^+^/CD45RA^-^ |
| **B cells** | B |  |
| Naïve B cells | Naïve B | CD3^-^CD19^+^/IgD^+^CD27^-^ |
| Unswitched memory B cells | USM B | CD3^-^CD19^+^/IgD^+^CD27^+^ |
| Switched memory B cells | SM B | CD3^-^CD19^+^/IgD^-^CD27^+^ |
| Double negative B cells | DN B | CD3^-^CD19^+^/IgD^-^CD27^-^ |
| Plasmablasts | Plasmablast | CD3^-^CD19^+^/IgD^-^/CD27^++^CD38^+^ |
| **Natural killer cells** | NK | CD3^-^CD19-/CD14-/CD56+ |
| **Monocytes** | Mono |  |
| CD16 positive monocytes | CD16p Mono | CD3^-^CD19^-^/HLADR^+^/CD56^-^/CD14^+^CD16^+^ |
| CD16 negative monocytes | CD16n Mono | CD3^-^CD19^-^/HLADR^+^/CD56^-^/CD14^+^CD16^-^ |
| **Dendric cells** | DC |  |
| Myeloid dendritic cells | mDC | CD3^-^CD19^-^/CD56^-^CD16^-^/HLADR^+^CD14^-^/CD11c^+^CD123^-^ |
| Plasmacytoid dendritic cells | pDC | CD3^-^CD19^-^/CD56^-^CD16^-^/HLADR^+^CD14^-^/CD11c^-^CD123^+^ |
| **Neutrophils** | Neu | Immune-magnetically sorting with the "EasySep direct human neutrophil isolation kit" |

**Online supplementary table 11. Definitions of the peripheral blood immune cell subsets in Dataset 4.**

| Dataset 4 |  |  |
| --- | --- | --- |
| Subset name | Abbreviation | Definition |
| **CD4 T cells** | CD4 |  |
| Naïve CD4 T cells | Naïve CD4 | CD3^+^/CD4^+^CD8^-^/CCR7+CD45RA+ |
| Memory CD4 T cells | Mem CD4 | CD3^+^/CD4^+^CD8^-^/non-naive CD4^+^/CD25^-^ |
| T helper 1 cells | Th1 | CD3^+^/CD4^+^CD8^-^/non-naive CD4^+^/CD25^-^/CXCR5^-^CCR6^-^/CXCR3^+^CCR4^-^ |
| T helper 2 cells | Th2 | CD3^+^/CD4^+^CD8^-^/non-naive CD4^+^/CD25^-^/CXCR5^-^CCR6^-^/CXCR3^-^CCR4^+^ |
| T helper 17 cells | Th17 | CD3^+^/CD4^+^CD8^-^/non-naive CD4^+^/CD25^-^/CXCR5^-^CCR6^+^/CXCR3^-^ |
| T follicular helper cells | Tfh | CD3^+^/CD4^+^CD8^-^/non-naive CD4^+^/CD25^-^/CXCR5^+^ |
| Fraction II effector  regulatory T cells | Fr. II eTreg | CD3^+^/CD4^+^CD8^-^/CD25^++^CD45RA^-^ |
| Fraction I naïve regulatory  T cells | Fr. I nTreg | CD3^+^/CD4^+^CD8^-^/CD25^+^CD45RA^+^ |
| Fraction III non-regulatory  T cells | Fr. III T | CD3^+^/CD4^+^CD8^-^/CD25^+^CD45RA^-^ |
| **CD8 T cells** | CD8 |  |
| Naïve CD8 T cells | Naïve CD8 | CD3^+^CD19^-^/CD4^-^CD8^+^/CD45RA^+^CCR7^+^ |
| CD8+ T effector memory  CD45RA+ cells | TEMRA CD8 | CD3^+^CD19^-^/CD4^-^CD8^+^/CD45RA^+^CCR7^-^ |
| Effector Memory CD8 T cells | EM CD8 | CD3^+^CD19^-^/CD4^-^CD8^+^/CD45RA^-^CCR7^-^ |
| Central Memory CD8 T cells | CM CD8 | CD3^+^CD19^-^/CD4^-^CD8^+^/CD45RA^-^CCR7^+^ |
| **B cells** | B |  |
| Naïve B cells | Naïve B | CD3^-^CD19^+^/IgD^+^CD27^-^ |
| Unswitched memory B cells | USM B | CD3^-^CD19^+^/IgD^+^CD27^+^ |
| Switched memory B cells | SM B | CD3^-^CD19^+^/IgD^-^CD27^+^/CD38^-^ |
| Double Negative B cells | DN B | CD3^-^CD19^+^/IgD^-^CD27^-^ |
| Plasmablasts | Plasmablast | CD3^-^CD19^+^/IgD^-^CD27^++^/CD38^+^ |
| **Natural Killer cells** | NK | CD3^-^CD19^-^/CD14^-^/CD56^+^ |
| **Monocytes** | Mono |  |
| CD16 positive monocytes | CD16p Mono | CD3^-^CD19^-^/HLA-DR^+^/CD56^-^/CD14^+^CD16^+^ |
| Non-classical monocytes | NC Mono | CD3^-^CD19^-^/HLA-DR^+^/CD56^-^/CD14^dim^CD16^+^ |
| Intermediate monocytes | Int Mono | CD3^-^CD19^-^/HLA-DR^+^/CD56^-^/CD14^++^CD16^+^ |
| CD16 negative monocytes | CD16n Mono | CD3^-^CD19^-^/HLA-DR^+^/CD56^-^/CD14^+^CD16^-^ |
| **Dendric cells** | DC |  |
| Myeloid dendritic cells | mDC | CD3^-^CD19^-^/HLA-DR^+^/CD56^-^/CD14^-^CD16^-^/CD11c^+^CD123^-^ |
| Plasmacytoid dendritic cells | pDC | CD3^-^CD19^-^/HLA-DR^+^/CD56^-^/CD14^-^CD16^-^/CD11c^-^CD123^+^ |
| **Neutrophils** | Neu | Immune-magnetically sorting with the "MACSxpress Neutrophil isolation Kit, human" |
| Low-Density Granulocytes | LDG | SSC^high^/CD16^+^CD15^+^/CD14^-^ |

**Online supplementary table 12. Fluorescently labelled antibodies used for PBMC staining in each Dataset.**

| Dataset 1-3 | |  |  |  | Dataset 4 | |  |  |
| --- | --- | --- | --- | --- | --- | --- | --- | --- |
| Panel 1 (for CD4 T cells) | | |  |  | Panel 1 (for CD4 T cells) | | |  |
| marker | conjugate | clones | company |  | marker | conjugate | clones | company |
| CD25 | AF488 | BC96 | eBioscience |  | CD25 | FITC | M-A251 | BD |
| CXCR5 | PerCP-Cy5.5 | RF8B2 | BD |  | CXCR5 | PerCP/Cy5.5 | RF8B2 | BD |
| CD3 | PE/Cy7 | UCHT1 | BioLegend |  | CD3 | PE/Cy7 | UCHT1 | BioLegend |
| CCR6 | APC | 11A9 | BD |  | CCR6 | APC | 11A9 | BD |
| CD45RA | APC/Cy7 | HI100 | BioLegend |  | CD45RA | APC/Cy7 | HI100 | BioLegend |
| CXCR3 | BV421 | 1C6/CXCR3 | BD |  | CXCR3 | BV421 | 1C6/CXCR3 | BD |
| CD4 | V500 | RPA-T4 | BioLegend |  | CD4 | BV510 | RPA-T4 | BD |
|  |  |  |  |  | CCR7 | BV711 | G043H7 | BioLegend |
|  |  |  |  |  | CCR10 | BUV395 | 1B5 | BD |
|  |  |  |  |  | CD8 | BUV805 | RPA-T8 | BD |
|  |  |  |  |  | CCR4 | BV605 | L291H4 | BioLegend |
| Panel 2 (for Mono, NK and DCs) | | |  |  | Panel 2 (for Mono, NK and DCs) | | |  |
| marker | conjugate | clones | company |  | marker | conjugate | clones | company |
| CD14 | FITC | M5E2 | BioLegend |  | CD14 | FITC | M5E2 | BioLegend |
| HLA-DR | PE | L243 | eBioscience |  | HLA-DR | PE | L243 | BioLegend |
| CD16 | PerCP/Cy5.5 | 3G8 | BioLegend |  | CD16 | PerCP/Cy5.5 | 3G8 | BioLegend |
| CD3 | PE/Cy7 | UCHT1 | BioLegend |  | CD3 | PE/Cy7 | UCHT1 | BioLegend |
| CD123 | APC | AC145 | Miltenyi |  | CD123 | APC | AC145 | Miltenyi Biotec |
| CD56 | APC/Cy7 | HCD56 | BioLegend |  | CD56 | APC/Cy7 | HCD56 | BioLegend |
| CD11c | BV421 | B-ly6 | BD |  | CD11c | BV421 | B-ly6 | BD |
| CD19 | V500 | HIB19 | BD |  | CD19 | BV510 | HIB19 | BD |
|  |  |  |  |  | CD141 | BV605 | 1A4 | BD |
|  |  |  |  |  | CD15 | BV711 | W6D3 | BD |
|  |  |  |  |  | CD1c | BUV395 | F10/21A3 | BD |
| Panel 3 (for B, CD8 T cells) | | |  |  | Panel 3 (for B, CD8 T cells) | | |  |
| marker | conjugate | clones | company |  | marker | conjugate | clones | company |
| CD27 | FITC | O323 | BioLegend |  | CD19 | BV605 | HIB19 | BioLegend |
| CD19 | PE | HIB19 | BioLegend |  | CD27 | FITC | O323 | BioLegend |
| CD38 | PerCP/Cy5.5 | HIT2 | BioLegend |  | CD38 | PerCP/Cy5.5 | HIT2 | BioLegend |
| CD3 | PE/Cy7 | UCHT1 | BioLegend |  | CD3 | PE/Cy7 | UCHT1 | BioLegend |
| CD45RA | APC | HI100 | BioLegend |  | CD45RA | APC | HI100 | BioLegend |
| CD4 | APC/Cy7 | RPA-T4 | BioLegend |  | CD4 | APC/Cy7 | RPA-T4 | BioLegend |
| IgD | BV421 | IA6-2 | BD |  | IgD | BV421 | IA6-2 | BD |
| CD8a | V500 | RPA-T8 | BD |  | CD8 | BV510 | RPA-T8 | BD |
|  |  |  |  |  | CCR7 | BV711 | G043H7 | BioLegend |

**Online supplementary table 13. Number of samples analyzed for each subset.**

| subset | HC | RA before treatment | RA after ABT treatment | RA after TCZ treatment |
| --- | --- | --- | --- | --- |
| Naïve CD4 | 34 | 49 | 14 | 4 |
| Mem CD4 | 35 | 50 | 15 | 4 |
| Th1 | 32 | 27 | 5 | 0 |
| Th2 | 27 | 27 | 5 | 0 |
| Th17 | 31 | 27 | 5 | 0 |
| Tfh | 32 | 28 | 5 | 0 |
| Fr. II eTreg | 37 | 49 | 15 | 5 |
| Naïve B | 34 | 54 | 15 | 5 |
| USM B | 34 | 51 | 15 | 5 |
| SM B | 35 | 50 | 15 | 5 |
| DN B | 35 | 51 | 14 | 5 |
| Plasmablast | 38 | 46 | 13 | 5 |
| NK | 36 | 49 | 15 | 4 |
| CD16p Mono | 38 | 52 | 15 | 5 |
| CD16n Mono | 37 | 52 | 15 | 3 |
| mDC | 37 | 45 | 13 | 4 |
| pDC | 36 | 45 | 13 | 5 |
| Neu | 34 | 45 | 13 | 3 |
| HC, healthy control; RA, rheumatoid arthritis. | | | | |

**Online supplementary table 14. Sequences of primer pairs used in qPCR.**

| gene# | Target | Sequence (Forward) | Sequence (Reverse) |
| --- | --- | --- | --- |
| 1 | COTL1 | 5'-TCGGTGGTGAGAAGGTTAGG-3' | 5'-GTGGGAACAGGTGGTTTCAG-3' |
| 2 | IFI30 | 5'-TACGGAAACGCACAGGAACA-3' | 5'-CAGGCCTCCACCTTGTTGAA-3' |
| 3 | FGL2 | 5'-TGTCCCAGCCAAGAACAAAT-3' | 5'-GCTTCTTTTGCCTATTGCGTA-3' |
| 4 | ABI3 | 5'-TAAGCACGCTGGGCCAGAT-3' | 5'-CAGCCAAAGTTGAGGGGTCT-3' |
| 5 | CX3CR1 | 5'-TCCTTCTGGTGGTCATCG-3' | 5'-TGTGCATTGGGTCCATCA-3' |
| 6 | LILRA2 | 5'-CACTTCCTGTGTGGTTGCAC-3' | 5'-GAGGGATTTTTCCCCTGAAG-3' |
| 7 | GFRA2 | 5'-CCGACTTCCATGCCA ATTGT-3' | 5'-ATGTCAAACCCAATCATGCCA-3' |
| 8 | KLF4 | 5'-CATCTCAAGGCACACCTGCGAA-3' | 5'-TCGGTCGCATTTTTGGCACTGG-3' |
| 9 | CYP2S1 | 5'-CGATGCCTTCCTGCTGAAG-3' | 5'-GCATGTTCTTGTTGGTGAATTCTG-3' |
| 10 | CD22 | 5'-CATCTCCTCGGCCCCTGGCT-3' | 5'-ATCCAGACGCAGGCCCCCTC-3' |
| 11 | TIMP1 | 5'-TCTGCAATTCCGACCTCGTC-3' | 5'-CTGTTCCAGGGAGCCACAAA-3' |
| 12 | S100A10 | 5'-GACCACACCCATATGCCATCT-3' | 5'-TATCAGGGAGGATCCAACTGC-3' |
| 13 | SLC24A4 | 5'-AGCAAGTCGGCTTCGTGT-3' | 5'-TGGGGCCATCAACTTTCTAT-3' |
| 14 | CEBPD | 5'-ACTCAGCAACGACCCATACC-3' | 5'-CGCTCCTATGTCCCAAGAAA-3' |
| 15 | NFE2 | 5'-ATCATGTCCATCACCGAGCTGC-3' | 5'-TGGGTCTTCTTGGGGCTTAGGT-3' |

**Online supplementary table 15. Stained antibodies used in the confirmatory cohort analyzed by mass cytometry.**

| marker | label | clones | company |
| --- | --- | --- | --- |
| CD279 (PD-1) | 174Yb | EH12.2H7 | FLUIDIGM |
| CD4 | 145Nd | RPA-T4 | FLUIDIGM |
| CD38 | 144Nd | HIT2 | FLUIDIGM |
| CD3 | 170Er | UCHT1 | FLUIDIGM |
| CD27 | 155Gd | L128 | FLUIDIGM |
| IgA | 148Nd | Polyclonal | FLUIDIGM |
| IgD | 146Nd | IA6-2 | FLUIDIGM |
| CD28 | 160Gd | CD28.2 | FLUIDIGM |
| CD223/LAG-3 | 165Ho | 11C3C65 | FLUIDIGM |
| CD49b | 161Dy | P1E6-C5 | FLUIDIGM |
| IgM | 172Yb | MHM-88 | FLUIDIGM |
| CD69 | 162Dy | FN50 | FLUIDIGM |
| CX3CR1 | 156Gd | 2A9-1 | FLUIDIGM |
| CD161 | 164Dy | HP-3G10 | FLUIDIGM |
| CD8a | 168Er | SK1 | FLUIDIGM |
| CD56 (NCAM) | 176Yb | NCAM16.2 | FLUIDIGM |
| CD45RA | 153Eu | HI100 | FLUIDIGM |
| CD134(OX40) | 158Gd | ACT35 | FLUIDIGM |
| CD25 (IL-2R) | 169Tm | 2A3 | FLUIDIGM |
| CD194/CCR4 | 149Sm | L291H4 | FLUIDIGM |
| CD197 (CCR7) | 167Er | G043H7 | FLUIDIGM |
| CD11c | 159Tb | Bu15 | FLUIDIGM |
| CD14 | 175Lu | M5E2 | FLUIDIGM |
| S1PR4 | 154Sm | 1 | AVIVA SYSTEMS BIOLOGY |
| CD183 (CXCR3) | 163Dy | G025H7 | FLUIDIGM |
| CD185 (CXCR5) | 171Yb | RF8B2 | FLUIDIGM |
| CD19 | 142Nd | HIB19 | FLUIDIGM |
| CD20 | 147Sm | 2H7 | FLUIDIGM |
| CD123 (IL-3R) | 151Eu | 6H6 | FLUIDIGM |
| CD16 | 209Bi | 3G8 | FLUIDIGM |
| CD196 (CCR6) | 141Pr | G034E3 | FLUIDIGM |
| CD63 | 150Nd | H5C6 | FLUIDIGM |
| CD24 | 166Er | ML5 | FLUIDIGM |
| CD39 | 152Sm | A1 | FLUIDIGM |
| CD278/ICOS | 143Nd | C398.4A | FLUIDIGM |
| HLA-DR | 173Yb | L243 | FLUIDIGM |

**Online supplementary table 16. Stained antibodies used in the pre-DC gating flow cytometry assessment.**

| marker | conjugate | clones | company |
| --- | --- | --- | --- |
| CD14 | FITC | M5E2 | BioLegend |
| HLA-DR | PE | L243 | BioLegend |
| CD16 | PerCP/Cy5.5 | 3G8 | BioLegend |
| CD3 | PE/Cy7 | UCHT1 | BioLegend |
| CD123 | APC | AC145 | Miltenyi Biotec |
| CD56 | APC/Cy7 | HCD56 | BioLegend |
| CD11c | BV421 | B-ly6 | BD |
| CD19 | BV510 | HIB19 | BD |
| CD141 | BV605 | 1A4 | BD |
| CD45RA | BV650 | HI100 | BD |
| CD33 | BV711 | WM53 | BD |
| CX3CR1 | BUV805 | 2A9-1 | BD |
| CD34 | BUV395 | 581 | BD |
